# Gut microbiome signatures of colorectal cancer development are more pronounced in women compared to men in a population-based screening cohort

**DOI:** 10.1101/2025.05.16.25327767

**Authors:** C. Bucher-Johannessen, AS. Kværner, E. Birkeland, E. Botteri, E. Avershina, V. Bemanian, G. Hoff, KR. Randel, E. Hovig, P. Berstad, TB. Rounge

## Abstract

**Background:** The gut microbiome has emerged as a promising source of biomarkers to enhance early detection of colorectal cancer (CRC). However, sex-specific differences in gut microbial profiles and their relationship to CRC risk remain underexplored.

**Objective:** To investigate sex-specific differences in gut microbial profiles of a CRC-screening population, as well as the potential for sex-specific associations between the gut microbiome and colorectal lesions.

**Methods:** This cross-sectional study included 1,034 faecal immunochemical test-positive screening participants aged 55-77 years recruited from the Norwegian CRCbiome study. Shotgun metagenomic sequencing was used to generate taxonomic and functional profiles of the gut microbiome, which were integrated with clinicopathological, demographic, and lifestyle data. Associations between sex, colorectal lesions, and microbial characteristics - including α-diversity, β-diversity, and abundances of bacterial species and functions - were assessed, including their interactions.

**Results:** Male participants had significantly higher odds of presenting with both non-advanced (OR: 1.50; 95% CI: 1.00-2.26) and advanced (OR: 1.46; 95% CI: 1.10-1.93) colorectal lesions compared to women. Gut microbial profiles differed markedly by sex, demonstrating compositional shifts and distinct bacterial profiles (13 bacteria and 41 functions more abundant in women, 19 taxa and 58 functions more abundant in men). In women, microbial α- and β-diversity varied across lesion subtypes, whereas no such differences were observed in men. Interaction analyses identified five bacteria and nine functions that were differentially associated with colorectal lesions by sex. Known CRC-associated bacteria showed broadly similar profiles in women and men, however, *pks*-positive *Escherichia coli* was associated with CRC in women only.

**Conclusion:** This study highlights sex-specific differences in the gut microbiome and their association with colorectal lesions, emphasising the need to take sex into account in future research aiming to enhance CRC prevention strategies and treatment.

**Trial Registration:** The BCSN is registered at clinicaltrials.gov (National clinical trial (NCT) no. 01538550).

## Introduction

Colorectal cancer (CRC) is the third most common cancer and the second leading cause of cancer-related deaths worldwide (1). Both CRC incidence and mortality show a male predominance, a pattern that remains consistent across countries, despite significant variations in incidence rates (2, 3). Variations in exposure to, and effects of, dietary and lifestyle factors are likely contributors to the observed disparities (4); however, the extent of their contribution remains unclear. Projections suggest that the observed sex disparities in CRC risk are likely to persist in the coming decades (2).

CRC screening has the potential to reduce both CRC incidence (5–8) and mortality (5–12). Faecal immunochemical testing (FIT) is the most widely used screening method (13), and is recommended as the preferred test by EU guidelines (14). FIT detects haemoglobin in stool, with positivity thresholds varying from >10 to >150 μg/g across European screening programs (15), prompting follow-up diagnostic colonoscopy. While FIT has a good sensitivity for colorectal cancers (16), it is less accurate for precancerous lesions (16) and lesions located in the proximal colon (17). Additionally, FIT-based screening has been criticised for potentially disadvantaging women (18). This may be due to anatomical, physiological, and/or pathogenetic differences affecting diagnostic performance of either the FIT (19–23) or the colonoscopy (24–27). Complementary strategies to improve screening efficacy and equity are needed.

Emerging evidence supports a role of the gut microbiome in CRC development (28–33). Proposed mechanisms include interaction with the immune system (34–36), induction of inflammation and oxidative stress (37–39), cross-talk with the endocrine system (40–42), and production of bioactive compounds, including metabolites, toxins, and virulence factors (43–49). A growing number of studies suggest that specific microbes — alone or in combination with other biomarkers — can be used to predict colorectal lesions (50, 51), and in a recent study, we showed that microbial signatures combined with quantitative FIT values can improve detection of premalignant lesions in a real-world screening setting (52). As sex impacts the microbial profile (53–56), it is plausible that the microbiome’s role in CRC development, as well as its potential as a biomarker source, varies by sex.

Our study provides an in-depth characterization of sex differences in gut microbial profiles and explores how these differences evolve with advancing CRC stage in a screening setting. Further, we examine whether the relationship between the gut microbiome and disease stage manifests differently in men and women, which is crucial for understanding the potential of the gut microbiome as a source of biomarkers in CRC screening.

## Methods and materials

### Study population

In this study, we used data from the CRCbiome study, which has been described in detail previously (57). The CRCbiome study was initiated in 2017 and invited participants from the large Bowel Cancer Screening in Norway pilot trial (BCSN) (n=139,291) (58). The BCSN started in 2012, where Norwegian citizens from selected municipalities surrounding the capital of Oslo (aged 50-74 years) were invited to either undergo once-only sigmoidoscopy or FIT test biennially. The CRCbiome study recruited participants from the FIT arm (rounds 2-4), using a positive FIT test as an inclusion criterion (≥15 µg haemoglobin/g faeces). Participants with a positive FIT test were referred to colonoscopy, where potential neoplastic lesions were removed. An information letter and two questionnaires, a lifestyle and demographic questionnaire (LDQ) and a food frequency questionnaire (FFQ), were sent out prior to scheduled colonoscopy. Participants were informed that by completing and returning either questionnaire, they provided consent and agreed to participate in the CRCbiome study. Samples were excluded from analyses if they had insufficient sequencing depth, defined as less than 1 gigabase of sequencing output after quality, if the individuals did not attend follow-up colonoscopy or presented with one or two non-advanced neoplastic lesions or serrated lesions during screening.

The BCSN trial and the CRCbiome study have been approved by the Regional Committee for Medical and Health-related Research Ethics in Southeast Norway (REK ref: 2011/1272 and 63148, respectively. The BCSN trial was registered at clinicaltrials.gov (Clinical Trial (NCT) no.: 01538550).

### Outcome measures

In this study, participants were classified into three colonoscopy groups: 1) “No neoplasia”, including individuals without lesions, those with ≥1 non-neoplastic lesion (e.g., inflammatory or granulation tissue polyps), and those with other findings (e.g., diverticulitis, haemorrhoids, or IBD); 2) “Non-advanced neoplastic lesions, and 3) “Advanced lesions”, which included those with ≥1 advanced serrated lesion, ≥1 advanced adenoma, or CRC. Here advanced adenoma was defined as any adenoma with at least one of the following characteristics: villous histology (≥25% villous components), high-grade dysplasia, or a polyp size ≥10 mm. Advanced serrated lesions were defined as serrated lesions with a size ≥10 mm or evidence of dysplasia. CRC was defined as the presence of adenocarcinoma originating in the colon or rectum (ICD-10 codes C18-20).

### Questionnaire data

Lifestyle and demographic information was collected using a self-administered four-page questionnaire, described in detail elsewhere (57). The questions relevant to the current study captured data on demographic factors (national affiliation, education, employment status, and marital status), clinical factors (family history of CRC and diagnosis of chronic bowel disorders), and lifestyle factors (smoking and snus habits, and physical activity level).

Dietary intake data were obtained using a validated 14-page semi-quantitative FFQ, developed and validated at the Department of Nutrition, University of Oslo (59–64). This FFQ assessed habitual dietary intake over the preceding year and provided estimates of total energy intake, macronutrient and micronutrient intake, as well as intake of foods and beverages (256 questions in total). Dietary intake in grams per day was calculated using the dietary calculation system KBS (short for “**K**ost**b**eregnings**s**ystem”). The most recent database at the time (AE-18) was used, which is an extended version of the 2018 Norwegian Food Composition Table (65). The FFQ also recorded self-reported body weight (kg) and height (cm), which were used to calculate body mass index (BMI). Based on data from the two questionnaires, a seven-point diet and lifestyle score was created to measure adherence to the 2018 World Cancer Research Fund (WCRF)/American Institute for Cancer Research (AICR) cancer prevention recommendations. The recommendations were operationalised using a standardised scoring system developed by Shams-White *et al*. (66, 67), as described in detail elsewhere (68).

### Prescription drug data

Dispensed prescription data within 12 months of study inclusion for each participant were retrieved from the Norwegian Prescription Database (NorPD), which catalogues all prescriptions in Norway using the Anatomical Therapeutic Chemical (ATC) system (69). The list included drugs for common comorbidities: diabetes (≥2 dispensations of any drug with ATC code A10); cardiovascular disease (≥1 of any with ATC codes C, and/or B01); chronic obstructive pulmonary disease (≥1 of any with ATC codes 698 R03AC, R03AK, R03AL, R03BB, R03DA, R03DC, R03DX, R03BA); and antibiotics (≥1 of any with 699 ATC codes J01, A07AA, or P01AB), since they are of relevance to microbiome analyses.

### Sample collection and DNA extraction

Participants were provided with at-home stool sampling kits suitable for FIT-measurement analyses and were instructed to ship the sample by mail to the testing facility (estimated delivery time 3-10 days). After FIT-measurement (testing for haemoglobin concentration), the leftover sample was further stored at -80°C. Before DNA extraction, leftover FIT samples were buffered, and 500 µL aliquots were used for analysis. DNA extraction utilised the QIAsymphony DSP Virus/Pathogen Midikit (Qiagen, Hilden, Germany), according to the instruction manual, with additional off-board lysis and bead-beating. Extracted DNA was buffered in 60 µl AVE buffer. DNA from FIT tubes showed no detectable cross-contamination and harboured diverse microbial communities (70, 71).

### Library preparation and sequencing

Following extraction, sample quality was measured on a NanoDrop 2000 spectrophotometer (Thermo Fisher Scientific, MA, USA) and DNA concentration using a Qubit Fluorometer (Thermo Fisher Scientific, MA, USA). Samples with a DNA concentration above 0.7 ng/µl were considered eligible for library preparation and sequencing. Library preparation was performed using the Nextera DNA Flex Library Prep Reference Guide according to the manual, with the modification of reducing the reaction volume to one-fourth of the reference. Sample pools were sequenced on the Illumina NovaSeq system (Illumina Inc., CA, USA), resulting in 151 bp paired-end reads. Library preparation and sequencing were performed at the FIMM Technology Centre in Helsinki, Finland.

### Microbiome characterisation

Sequencing reads were quality controlled using KneadData (v0.12) (http://huttenhower.sph.harvard.edu/kneaddata) with default settings to remove adapters, low-quality bases, and reads mapping to the human genome. MetaPhlAn4 (72) (v4.0.6) was used to identify the relative abundance of microbial taxa and bacterial taxa for use in subsequent analyses. Further, HUMAnN3 was used (73) (v3.7) to determine gene content using the mpa_vOct22_ChocoPhlAn_202212 pangenome database and the UniRef90 (74) database for classifying the gene families into Gene Ontology (GO) annotations (75, 76) covering biological processes (BP), molecular functions (MF) and cellular components (CC) – hereafter collectively referred to as functions. Abundance of gene content from HUMAnN3 was scaled by the number of quality-assessed reads per million.

Targeted analyses were carried out for two sets of microbial constituents: Bacterial species identified as associated with the presence of CRC both by us (52) and others (77), and *pks*-stratified *E. coli*. To determine *pks* status, reads were aligned to the *pks* gene cluster (GenBank: AM229678.1). Samples with MetaPhlAn4-detected *E. coli* and ≥10% coverage of the *pks* region were classified as *pks*-positive, unless mapping showed an incomplete colibactin cluster.

## Statistics

We summarised continuous variables using the median value with interquartile range and categorical variables using counts with percentages. Within-group differences were examined using Kruskal-Wallis or Wilcoxon rank sum test for continuous variables and Pearson’s Chi-squared or Fisher’s exact test for categorical variables, as appropriate.

Missing data resulted from participants not returning or fully completing the questionnaires, or whose responses were of insufficient quality. Therefore, to ensure complete datasets, multivariate imputation by chained equations was performed using the R package *mice* (78). The imputed variables included marital status, employment status, smoking status, and the WCRF/AICR score. All variables listed in Table 1 were used as predictors in the imputation process. All analyses reported in the manuscript are based on the imputed dataset.

**Table 1.**
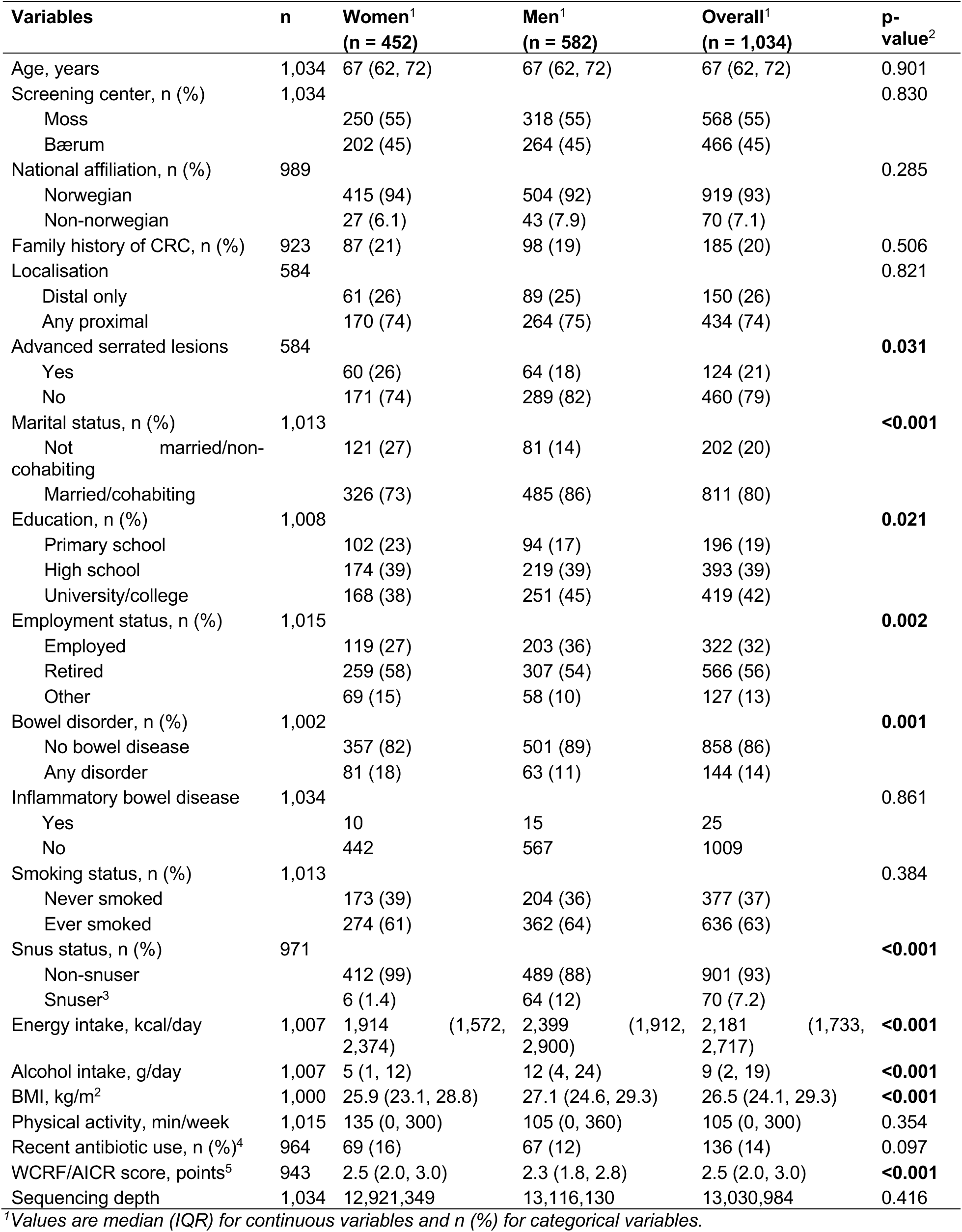
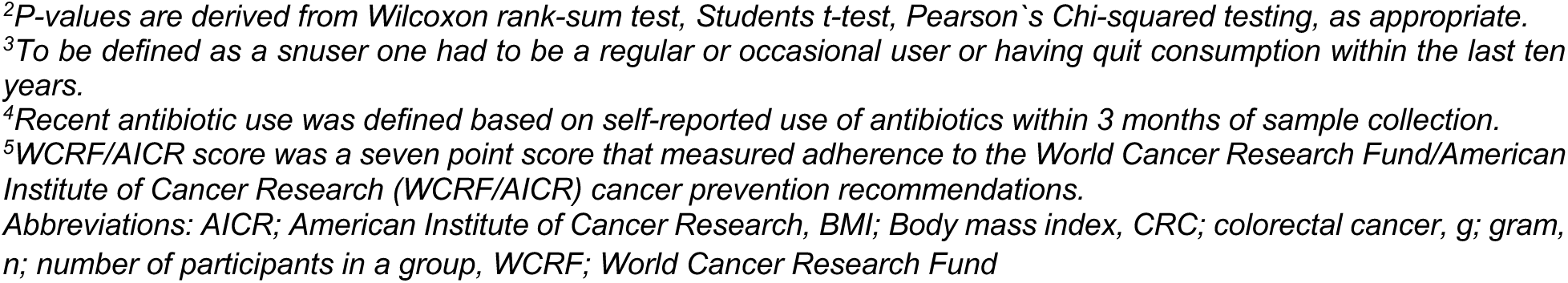
Participants’ characteristics overall (n=1,034) and among women (n=452) and men (n=582)^1^.

| Variables | n | Women<sup>1</sup><br>(n = 452) | Men<sup>1</sup><br>(n = 582) | Overall<sup>1</sup><br>(n = 1,034) | p-value<sup>2</sup> |
| --- | --- | --- | --- | --- | --- |
| Age, years | 1,034 | 67 (62, 72) | 67 (62, 72) | 67 (62, 72) | 0.901 |
| Screening center, n (%) | 1,034 |  |  |  | 0.830 |
| Moss |  | 250 (55) | 318 (55) | 568 (55) |  |
| Bærum |  | 202 (45) | 264 (45) | 466 (45) |  |
| National affiliation, n (%) | 989 |  |  |  | 0.285 |
| Norwegian |  | 415 (94) | 504 (92) | 919 (93) |  |
| Non-norwegian |  | 27 (6.1) | 43 (7.9) | 70 (7.1) |  |
| Family history of CRC, n (%) | 923 | 87 (21) | 98 (19) | 185 (20) | 0.506 |
| Localisation | 584 |  |  |  | 0.821 |
| Distal only |  | 61 (26) | 89 (25) | 150 (26) |  |
| Any proximal |  | 170 (74) | 264 (75) | 434 (74) |  |
| Advanced serrated lesions | 584 |  |  |  | **0.031** |
| Yes |  | 60 (26) | 64 (18) | 124 (21) |  |
| No |  | 171 (74) | 289 (82) | 460 (79) |  |
| Marital status, n (%) | 1,013 |  |  |  | **<0.001** |
| Not married/non-cohabiting |  | 121 (27) | 81 (14) | 202 (20) |  |
| Married/cohabiting |  | 326 (73) | 485 (86) | 811 (80) |  |
| Education, n (%) | 1,008 |  |  |  | **0.021** |
| Primary school |  | 102 (23) | 94 (17) | 196 (19) |  |
| High school |  | 174 (39) | 219 (39) | 393 (39) |  |
| University/college |  | 168 (38) | 251 (45) | 419 (42) |  |
| Employment status, n (%) | 1,015 |  |  |  | **0.002** |
| Employed |  | 119 (27) | 203 (36) | 322 (32) |  |
| Retired |  | 259 (58) | 307 (54) | 566 (56) |  |
| Other |  | 69 (15) | 58 (10) | 127 (13) |  |
| Bowel disorder, n (%) | 1,002 |  |  |  | **0.001** |
| No bowel disease |  | 357 (82) | 501 (89) | 858 (86) |  |
| Any disorder |  | 81 (18) | 63 (11) | 144 (14) |  |
| Inflammatory bowel disease | 1,034 |  |  |  | 0.861 |
| Yes |  | 10 | 15 | 25 |  |
| No |  | 442 | 567 | 1009 |  |
| Smoking status, n (%) | 1,013 |  |  |  | 0.384 |
| Never smoked |  | 173 (39) | 204 (36) | 377 (37) |  |
| Ever smoked |  | 274 (61) | 362 (64) | 636 (63) |  |
| Snus status, n (%) | 971 |  |  |  | **<0.001** |
| Non-snuser |  | 412 (99) | 489 (88) | 901 (93) |  |
| Snuser<sup>3</sup> |  | 6 (1.4) | 64 (12) | 70 (7.2) |  |
| Energy intake, kcal/day | 1,007 | 1,914 (1,572, 2,374) | 2,399 (1,912, 2,900) | 2,181 (1,733, 2,717) | **<0.001** |
| Alcohol intake, g/day | 1,007 | 5 (1, 12) | 12 (4, 24) | 9 (2, 19) | **<0.001** |
| BMI, kg/m<sup>2</sup> | 1,000 | 25.9 (23.1, 28.8) | 27.1 (24.6, 29.3) | 26.5 (24.1, 29.3) | **<0.001** |
| Physical activity, min/week | 1,015 | 135 (0, 300) | 105 (0, 360) | 105 (0, 300) | 0.354 |
| Recent antibiotic use, n (%)<sup>4</sup> | 964 | 69 (16) | 67 (12) | 136 (14) | 0.097 |
| WCRF/AICR score, points<sup>5</sup> | 943 | 2.5 (2.0, 3.0) | 2.3 (1.8, 2.8) | 2.5 (2.0, 3.0) | **<0.001** |
| Sequencing depth | 1,034 | 12,921,349 | 13,116,130 | 13,030,984 | 0.416 |

<sup>1</sup>Values are median (IQR) for continuous variables and n (%) for categorical variables.
<sup>2</sup>P-values are derived from Wilcoxon rank-sum test, Students t-test, Pearson's Chi-squared testing, as appropriate. <sup>3</sup>To be defined as a snuser one had to be a regular or occasional user or having quit consumption within the last ten years. <sup>4</sup>Recent antibiotic use was defined based on self-reported use of antibiotics within 3 months of sample collection. <sup>5</sup>WCRF/AICR score was a seven point score that measured adherence to the World Cancer Research Fund/American Institute of Cancer Research (WCRF/AICR) cancer prevention recommendations. Abbreviations: AICR; American Institute of Cancer Research, BMI; Body mass index, CRC; colorectal cancer, g; gram, n; number of participants in a group, WCRF; World Cancer Research Fund

We ran a multinomial logistic regression analysis to study the association between sex and colonoscopy findings (i.e., a) no neoplasia (reference category); b) non-advanced neoplastic lesions; and c) advanced lesions). The analysis was adjusted for the following covariates: Model 1) age (continuous) and screening centre (centre 1, centre 2); model 2: model 1 covariates, employment status (employed, retired, other), marital status (not married/non-cohabiting, married/cohabiting), smoking status (never smoked, ever smoked) and the WCRF/AICR score (continuous). Covariates were selected based on the association patterns observed in Table 1 and Supplementary Table 1. Unless otherwise specified, the subsequent analyses were conducted using these covariate sets.

To investigate microbial differences between sexes, α-diversity, β-diversity, and differential abundance of bacterial species and functions were assessed. Differences in α-diversity indices (Shannon and Inverse Simpson) were analysed using analysis of variance (ANOVA). β-diversity differences were assessed using the Bray-Curtis dissimilarity index, calculated with the vegdist function in the R package *vegan* (79) and tested for significance using permutational analysis of variance (PERMANOVA) with 999 permutations. Differentially abundant taxa and functions were identified using *MaAsLin2* (80) with the following settings: a minimum abundance threshold of 10%; log2 transformation with zero values replaced by a pseudo-count equal to half the lowest observed value across all features; normalisation set to none for bacterial abundances and total sum scaling for functions; and a linear model. Prior to analysis, functions strongly correlated with bacterial species (Spearman’s correlation coefficient > 0.5) were excluded to mitigate the risk of spurious associations driven by bacterial variation. Differentially abundant species and functions were adjusted for multiple testing using the Benjamini-Hochberg (BH) correction method.

To investigate sex-specific influences of the gut microbiome on colorectal lesions, sex-stratified analyses were conducted for α-diversity, β-diversity, differential abundance of microbial species and functions, presence or absence of CRC-associated species, and pks-stratified *E. coli*. For α-and β-diversity, sex-specific associations were examined by including interaction terms for sex and lesion status. Bacterial species and functions that were differentially abundant between no neoplasia and advanced lesions at a BH-corrected p-value of 0.2, in either men or women, were further analysed for interaction effects in the entire dataset using multinomial logistic regression with interaction terms of sex and the log-transformed abundance values. Evaluation of CRC-associated species was performed by sex-stratified logistic regression models contrasting no neoplasia with CRC, using the presence or absence of each CRC-associated species as the independent variable. Any *E. coli* detection and p*ks-*stratified *E. coli* was evaluated in a multinomial logistic regression model contrasting no neoplasia with non-advanced neoplastic lesions and advanced lesions, and a separate logistic regression model contrasting no neoplasia with CRC, performed independently in each sex.

To address potential sex-related differences in lesion subtype and location, sex-stratified α- and β-diversity analyses were performed comparing individuals with advanced serrated lesions to those without neoplasia, and by lesion localisation. Sensitivity analyses were also performed without including individuals with: advanced serrated lesions, inflammatory bowel disease (IBD) diagnosed during colonoscopy, and self-reported consumption of antibiotics within the last 3 months prior to study inclusion.

All analyses were performed using the R statistical environment with the main packages *tidyverse* (81) (v1.3.1), *rstatix* (82) (v0.7.0), *nnet* (83) (v7.3-16), *mice* (78) (v3.14.0), *vegan* (79) (v2.5.7), and *MaAsLin2* (80) (v1.12.0).

## Results

### Characteristics of the study population overall and by sex

Participants’ characteristics overall and stratified by sex are presented in Table 1. The study included more men (n=582; 56%) than women (n=452; 44%). Men were more likely to be married, possess a university degree, and be employed, whereas women more frequently reported bowel disorders. In terms of lifestyle factors, men were more likely to use tobacco snus, have a higher BMI, and lower adherence to cancer prevention recommendations compared to women. Similar patterns were also observed in the control group alone (Supplementary Table 1). No difference in lesion location (proximal versus distal) was observed between men and women. However, there was a significant difference in lesion subtypes, women more often presenting with advanced serrated lesions than men. In both sexes, the presence of screen-detected lesions (whether non-advanced or advanced) was associated with older age, being retired, being unmarried or not cohabiting, being screened at Bærum Hospital and having a higher alcohol consumption (Supplementary Table 2). Differences in comorbidity-related prescriptions across colonoscopy groups were seen for haemorrhoid and cardiovascular medications in men, and for type 2 diabetes mellitus (T2DM) drugs in women (Supplementary Table 3). In both sexes, individuals without neoplasia more frequently reported IBD (Supplementary Table 3). Supplementary Table 4 shows the associations between sex and colonoscopy findings. Compared to women, men were more likely to present with non-advanced neoplastic lesions and advanced lesions, with odds ratios (ORs) of 1.55 (95% confidence interval (CI): 1.05, 2.29) and 1.47 (95% CI: 1.12, 1.92), respectively. Additional adjustment for lifestyle and demographic factors only modestly attenuated the effect estimates (ORs of 1.50 (95% CI: 1.00, 2.26) and 1.46 (95% CI: 1.10, 1.93), respectively).

### Sex and the gut microbiome

Among the 1,034 included participants, the mean (SD) number of paired-end reads was 13,030,984 (3,883,470). A total of 2,756 bacterial species were identified, with a mean (SD) of 239 (68) species per sample, with means (SD) of 241 (66) and 237 (69) in men and women, respectively.

#### α- and β-diversity

Associations of sex with α- and β-diversity are illustrated in Figure 1A-B. There were no associations between sex and the two α-diversity indices (neither Shannon nor the Inverse Simpson index, ANOVA P > 0.05, Figure 1A). However, notable differences in β-diversity were observed between the sexes both in the control group (PERMANOVA, P = 0.002, R2 = 0.004, Supplementary Figure 1) and the population as a whole (PERMANOVA, P = 0.001, R2 = 0.003, Figure 1B), which were attenuated, but remained statistically significant after adjusting for lifestyle and demographic factors (PERMANOVA, P = 0.001, R2 = 0.002).

**Figure 1:**
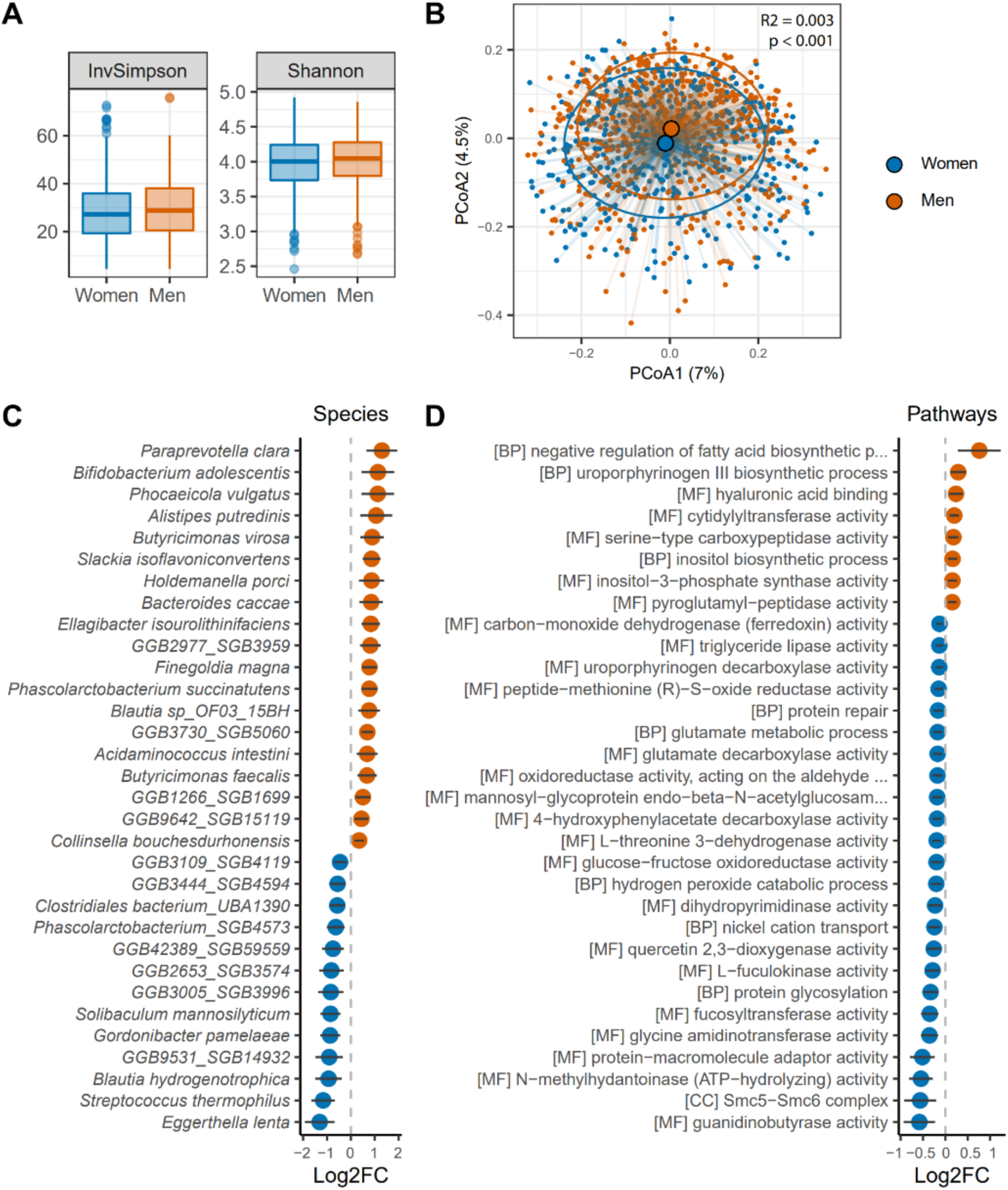
Sex-related differences in gut microbial profiles. Within-sample α-diversity, measured by the Inverse Simpson (left) and Shannon (right) indices, in women and men. (B) Between-sample β-diversity visualized by principal coordinates analysis (PCoA) of Bray–Curtis dissimilarity. Differences between sexes were assessed using PERMANOVA; corresponding R² values and p-values are shown in the plot. (C–D) Differentially abundant bacterial species (C) and microbial functions (D) between women and men identified by MaAsLin2 analyses. Effect estimates are shown as log₂ fold-change (Log₂FC) with 95% confidence intervals (CI). Species were included if Benjamini–Hochberg adjusted p-values were < 0.05; for microbial functions, the 32 features with the most extreme Log₂FC in either direction are displayed (full results in Supplementary Data 1). Features enriched in men are shown in red; those enriched in women are shown in blue. All models were adjusted for age (continuous), screening centre (Moss, Bærum), employment status (employed, retired, other), marital status (married/cohabiting, not married/non-cohabiting), smoking status (never, ever), and the continuous WCRF/AICR score. Names of the following functions have been truncated in D): [BP] negative regulation of fatty acid biosynthetic process; [MF] oxidoreductase activity, acting on the aldehyde or oxo group of donors, disulfide as acceptor; [MF] mannosyl-glycoprotein endo-beta-N-acetylglucosaminidase activity. Abbreviations: BP; biological process, CC; Cellular Components, MF; Molecular Function.

#### Differentially abundant bacteria and functions

In total, 52 bacterial species were differentially abundant between women and men (P*adj* < 0.05). Of these, 32 (62%) species remained statistically significant after adjusting for lifestyle and demographic factors (Figure 1C, Supplementary Data 1). Thirteen were more abundant in women and 19 in men. The strongest associations with male sex were observed for *Paraprevotella clara* and *Bifidobacterium adolescentis*. In contrast, the species most strongly associated with female sex were *Eggerthella lenta* and *Streptococcus thermophilus*. At the functional level, 174 functions were differentially abundant between the sexes (P*adj* < 0.05). Of these, 99 (57%) remained statistically significant after adjusting for lifestyle and demographic factors (Figure 1D, Supplementary Data 1), with 41 functions more abundant in women and 58 in men. In women, functions with the largest log2 fold-change (Log2FC) were guanidinobutyrase activity (MF), Smc5-Smc6 complex (CC), and N-methylhydantoinase (ATP-hydrolyzing) activity (MF). In men, the functions with the largest effect sizes were negative regulation of fatty acid biosynthetic processes (BP), uroporphyrinogen III biosynthetic processes (BP) and hyaluronic acid binding (MF).

#### Potential interactions between sex, the gut microbiome, and colorectal lesions

To evaluate whether the association between the gut microbiome and colorectal lesions differed by sex, sex-stratified analyses were performed for the following microbial measures: 1) α-diversity, 2) β-diversity, and 3) taxonomic and functional profiles. For all measures, notable sex differences were observed. Among women, significant differences in both α- and β-diversity were identified between colonoscopy groups (Figure 2A-B). Women with non-advanced neoplastic lesions had lower α-diversity compared to those without findings, while women with advanced lesions had higher α-diversity (ANOVA, P < 0.05 for all comparisons, Figure 2A). Additionally, distinct separation among colonoscopy groups was observed in the PCoA plot (PERMANOVA, *P* = 0.007, R² = 0.006, Figure 2B). Conversely, no significant differences in α- or β-diversity were observed across colonoscopy groups in men (Figure 2A and C). When examining advanced lesions by location (no lesion vs. proximal, distal, or both), women showed a clear but non-significant separation between groups, particularly between no lesion and distal lesions. This pattern was less pronounced in men (Supplementary Figure 2); however, combined analyses showed no statistically significant interaction effect (P = 0.066, P = 0.171, and P = 0.530, for the Shannon, Inverse Simpson, and Bray-Curtis index, respectively). Sensitivity analyses excluding participants with serrated lesions, IBD diagnosed during colonoscopy examination, or self-reported antibiotic use within 3 months of colonoscopy yielded similar results to the overall analyses.

**Figure 2.**
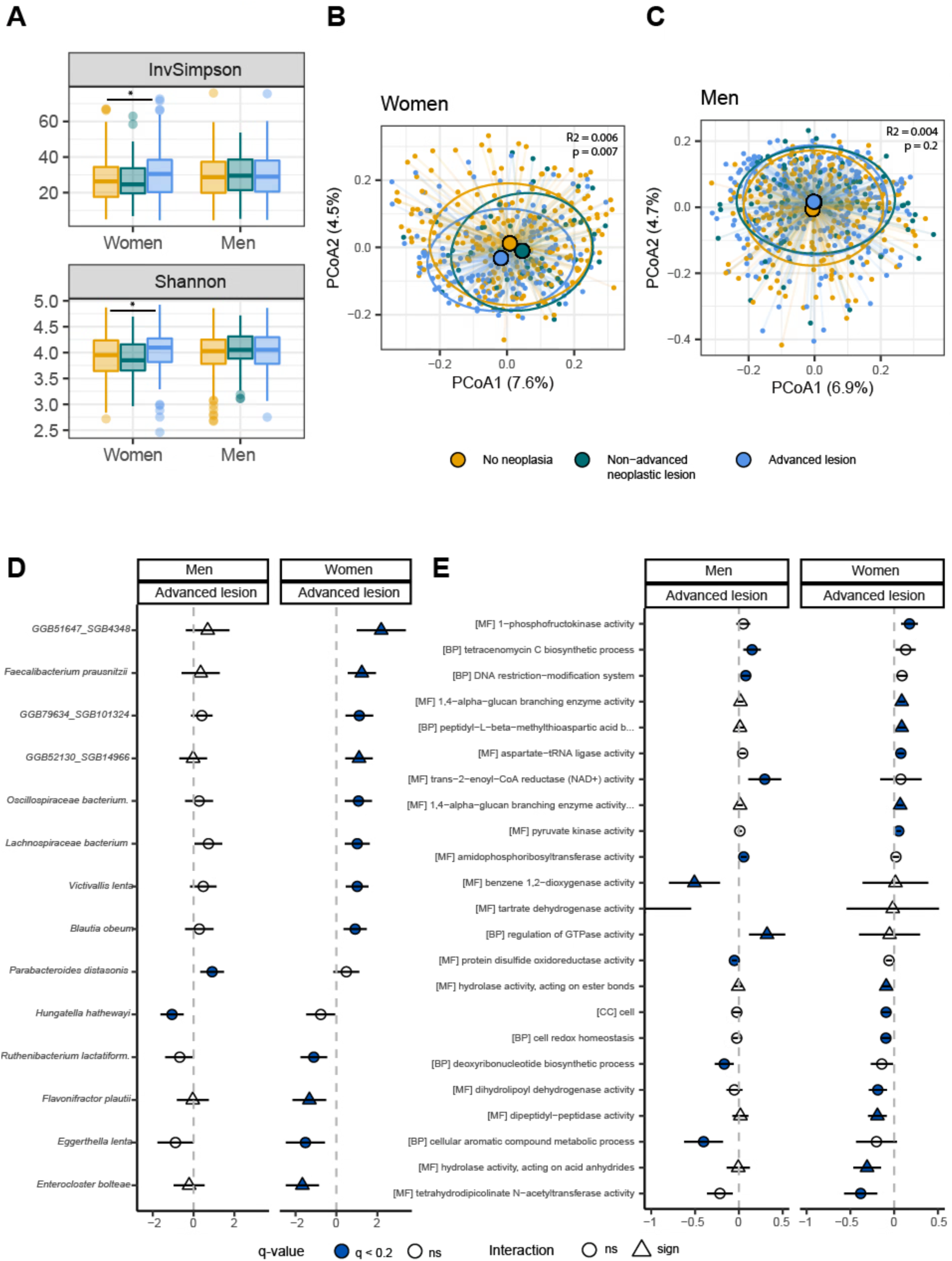
Sex-stratified analyses of gut microbial profiles across colonoscopy findings. (A) Within-sample α-diversity, measured by the Inverse Simpson (upper) and Shannon (lower) indices, across colonoscopy groups—no neoplasia (NN), non-advanced neoplastic lesions (NANL), and advanced colorectal lesions (AL)—in women and men. (B–C) Between-sample β-diversity visualized by PCoA of Bray–Curtis dissimilarity for women (B) and men (C). (D–E) Differentially abundant bacterial species (D) and microbial functions (E) across colonoscopy groups in women and men. Effect estimates comparing AL to NN are shown as Log₂FC with 95% CI. Species and functions were selected for visualization based on Benjamini–Hochberg adjusted p-values < 0.2 and < 0.15, respectively, in either sex (all features with p < 0.2 are listed in Supplementary Data 2). Triangles denote significant interactions between sex and abundance of species/functions identified by multinomial logistic regression (p < 0.05). All models were adjusted for age (continuous), screening centre (Moss, Bærum), employment status (employed, retired, other), marital status (married/cohabiting, not married/non-cohabiting), smoking status (never, ever), and the continuous WCRF/AICR score. Names of the following species and functions have been truncated in D): *Oscillospiraceae bacterium Marseille-Q3528* and E): 1,4-alpha-glucan branching enzyme activity (using a glucosylated glycogenin as primer for glycogen synthesis); peptidyl-L-beta-methylthioaspartic acid biosynthetic process from peptidyl-aspartic acid. Abbreviations: sign; significant, ns; non-significant, BP; biological process, CC; Cellular Components, MF; Molecular Function.

For bacterial and functional abundance, a sex-specific pattern emerged when comparing no neoplasia and advanced lesions. In women, 12 bacteria and 39 functions were differentially abundant between groups, whereas 2 bacteria and 18 functions were differentially abundant in men (P*adj* < 0.2, Figure 2D, Supplementary Data 2). Among the identified species, five (*GGB52130 SGB14966*, *Enterocloster bolteae, Faecalibacterium prausnitzii, GGB51647 SGB4348,* and *Flavonifractor plautii*) were confirmed to be sex-specific in multinomial logistic regression analyses (interaction terms P < 0.05). Notably, *E. lenta* and *Hungatella hathewayi* were inversely associated with advanced lesions regardless of sex. Among the differentially abundant functions, nine were confirmed to be sex-specific in multinomial logistic regression analyses (interaction terms P < 0.05), including two related to biological processes (peptidyl-L-beta-methylthioaspartic acid biosynthetic process from peptidyl-aspartic acid, regulation of GTPase activity), and seven related to molecular functions related to glycogen biosynthesis (1,4-alpha-glucan branching enzyme activity, 1,4-alpha-glucan branching enzyme activity (using a glycosylated glycogenin as primer for glycogen synthesis)), and small molecule degradation (benzene 1,2-dioxygenase activity, tartrate dehydrogenase activity, hydrolase activity, acting on ester bonds, dipeptidyl-peptidase activity, and hydrolase activity, acting on acid anhydrides).

#### Evaluation of previously reported CRC-associated species in women and men

When evaluating the presence or absence of previously CRC-associated (77) and validated (52) bacterial species (n = 9) in sex-stratified models (Figure 3A, Supplementary Table 5), effect estimates were broadly similar between men and women. All species but one (*F. nucleatum*) were significantly associated with CRC in at least one sex-stratified analysis. For women, *Gemella morbirollum*, *Peptostreptococcus stomatis,* and one SGB belonging to *F. nucleatum* was significantly associated with CRC. In men, *Dialister pneumosintes*, the uncharacterized SGB14305, *Alistipes ihumii*, *Porphyromonas somerae*, and *Porphyromonas uenonis* were significantly associated with CRC. In women, the prevalence of three CRC-associated species was very low; an overview of the prevalence of all the bacteria in men and women separately can be found in Supplementary Table 5. In a recent analysis, we identified *E. coli* as being associated with colorectal lesions in a *pks*-dependent manner (52). In analyses of *E. coli* detection (Figure 3B), the direction and magnitude of effect estimates were largely consistent between sexes, but significance levels differed. In women, *pks*-negative *E. coli* was significantly associated with non-advanced neoplastic lesions, and *pks*-positive *E. coli* was significantly associated with CRC. In men, both overall *E. coli* detection and *pks*-negative *E. coli* were significantly associated with non-advanced neoplastic lesions and advanced lesions. No significant interaction between sex and *E. coli* status was observed (interaction term P > 0.05 for all comparisons)

**Figure 3:**
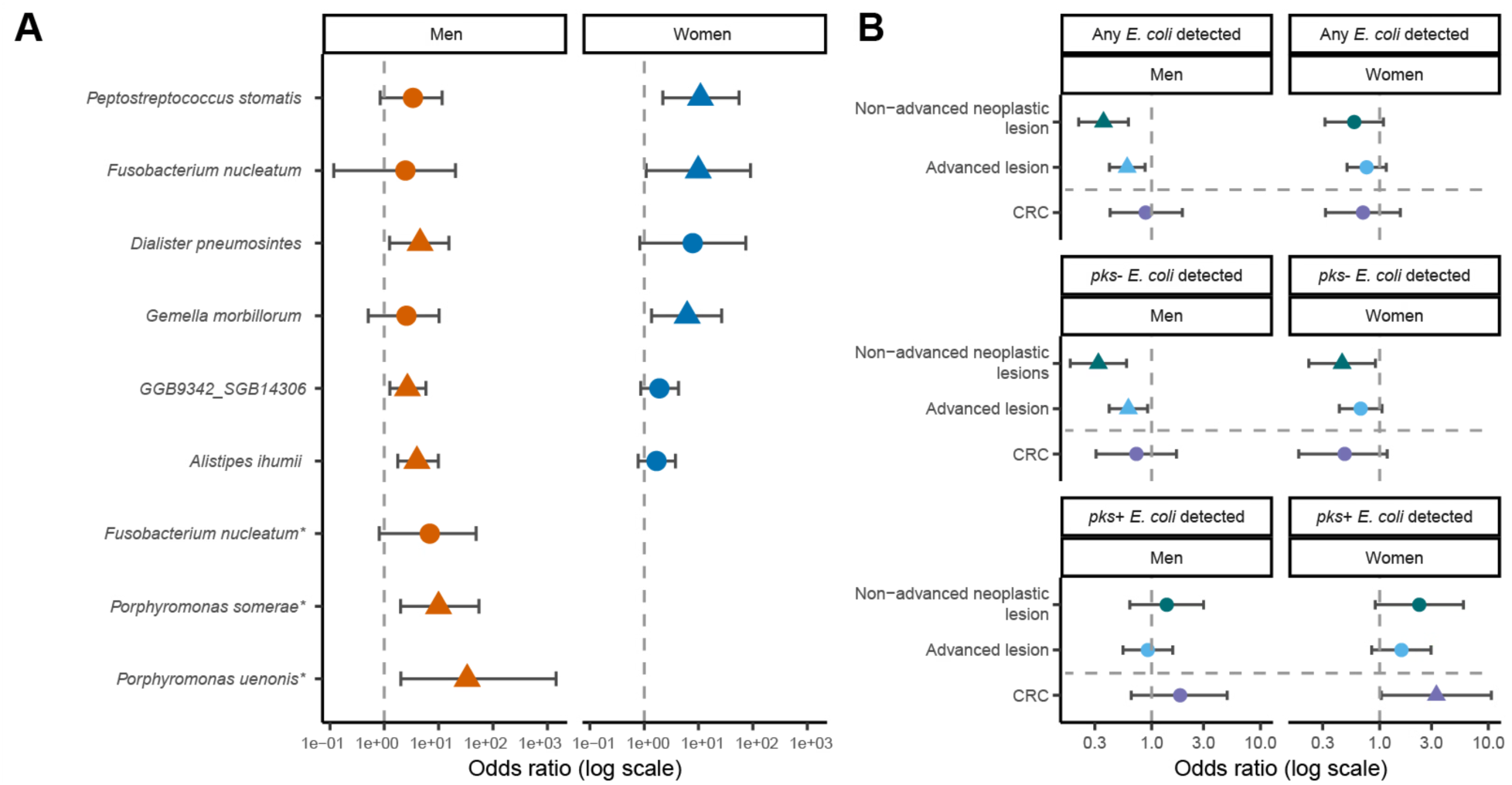
Sex-stratified associations of previously CRC-linked bacteria including *E. coli* detection and *pks*-status. (A) Odds ratios (log scale) from sex-stratified logistic regression models comparing participants with no neoplasia to those with CRC for previously reported CRC-associated bacterial species (77) validated in CRCbiome (52). Triangles denote associations significant at *P* < 0.05. *Odds ratio and CI are not displayed for women due to low prevalence across groups. (B) Odds ratio from regression models assessing associations between *E. coli* detection status (any *E. coli* (top), *pks*-negative *E. coli* (middle), and *pks*-positive *E. coli* (bottom) and lesion type (logistic regression for non-advanced neoplastic lesions, advanced lesion, and a multinomial logistic regression for CRC), shown separately for men and women. The reference group was participants with no neoplasia. Triangles denote associations significant at *P* < 0.05. All models were adjusted for age (continuous), screening centre (Moss, Bærum), employment status (employed, retired, other), marital status (married/cohabiting, not married/non-cohabiting), smoking status (never, ever), and the continuous WCRF/AICR score.

## Discussion

Recent work from our group, conducted in a large CRC screening-relevant cohort, demonstrated that microbiome signatures add a modest but significant increase in prediction value for detecting colorectal lesions (52), similar to other studies (48, 84–86). Yet, despite growing evidence that host factors influence the microbiome, especially in disease, sex-specific differences in the gut microbiome have remained largely unexplored. In this study, including metagenomes from >1000 CRC screening participants, we show that sex may serve as an effect modifier, where microbial associations with advanced colorectal lesions were stronger in women than in men. Still, CRC-associated bacteria did not show evidence of being sex-specific.

The CRCbiome study, including FIT-positive participants recruited from the large Norwegian colorectal screening trial BCSN, showed that men had a significantly higher odds of presenting with both non-advanced neoplastic lesions (55%) and advanced lesions (47%) at follow-up colonoscopy, consistent with previous reports (21), including results from BCSN (15, 58). The presence of advanced lesions is strongly linked to modifiable risk factors (87–89), as also documented in our cohorts (68, 90–93). It has been suggested that the association between male sex and CRC risk, at least in part, is caused by these factors (40, 94). However, the current study and those from a large German screening cohort (n=15,985(95)) suggest that a large part of the excess risk of advanced neoplasia among men cannot be explained by known modifiable risk factors and that these alone are insufficient to explain the sex gap in CRC risk. On the other hand, the higher occurrence of lesions in men could partially be attributed to FIT being less efficient in women. Therefore, sex-specific FIT thresholds have been suggested (23), but adjusting FIT thresholds is not straightforward (20, 21, 23, 96), highlighting the need for complementary strategies

Similarly, we show that disparities in microbial profile between men and women are not solely attributable to differences in exposure profiles. After adjusting for key lifestyle and demographic factors known to contribute to microbial variation, a large part of the sex-microbiome associations remained, suggesting that intrinsic factors are influential. Hormonal influence on the gut microbiome is supported by evidence showing that microbial differences between sexes appear in young adults and decline after age 50 (97). Further, oestrogens are known to exhibit anti-inflammatory properties, potentially modulating the gut environment and influencing cancer risk differently in men and women, whereas androgens seem to exacerbate cancer development (40). Indeed, the role of sex hormones in CRC development has been demonstrated in mice (98, 99). Distinct sex-specific microbial profiles have been reported (56, 115–118), including investigations within CRCbiome that have linked alcohol consumption to microbial variation by sex (91). We have also previously reported that men in CRCbiome have a higher abundance of *P. succinatutens* (100) and have identified CRC associations for bacteria of the *Phascolarctobacterium* genus across populations (100–102). In the present study, we identified 32 bacterial species that were differentially abundant between men and women. Here, we found that *B. adolescentis* levels were higher in men. This species has previously been found to be elevated in older male CRC patients (103) and has been proposed as a probiotic due to its pro-androgenic activity (104). In line with previous reports, we observed a higher abundance of *E. lenta* and *G. pamelae* in women (97, 105). Both species potentially play a role in host physiology and hormonal homeostasis via corticoid conversion to progestin (106). *E. lenta* has also been reported as being increased in women with premature ovarian insufficiency (107). The hormone-related *E. lenta* was the only species identified as both sex-specific and linked to the presence of lesions.

When examining microbial profiles separately in men and women, we observed that women exhibited clear differences in both α- and β-diversity across colonoscopy groups and lesion localisation, whereas no such pattern was observed in men. Previous studies have suggested sex differences in microbial profiles across controls, non-advanced neoplastic lesions, and CRC, with *Liao* et al. (108) also reporting greater variation in women during CRC development. However, these studies have been limited to a small sample size (N < 90), low taxonomic resolution, or lack thorough description of participant inclusion (108–111). Therefore, to our knowledge, our study is the first large-scale investigation within a population-based screening setting to demonstrate sex-specific differences in the microbiome across colorectal lesion subtypes.

We observed nominally significant, sex-specific associations for several bacterial species and microbial functions. Five species, including *F. plautii*, *E. bolteae*, and *F. prausnitzii*, were associated with advanced lesions in a sex-specific manner, with the effect size in directions opposite to those reported previously (77). In our recent work (52), we proposed that these taxa may reflect alternative causes of intestinal bleeding rather than being lesion-specific. One study found that older age, female sex, smoking, alcohol consumption, higher BMI, and haemorrhoids were associated with increased odds of false-positive FIT results in a screening population (112). However, such factors, including other lifestyle factors and comorbidity profiles, were comparable between sexes in our cohort, apart from a higher percentage of women with no neoplasia having a prescription for medication indicating type 2 diabetes mellitus (T2DM). Although T2DM and diabetes medication can influence the microbiome (113), the number of female participants with prescriptions for diabetes medication in this cohort remains small. Together, these observations suggest that intrinsic biological differences rather than external confounders are the primary drivers of the sex-specific microbiome patterns we observed.

We have previously verified and reproduced the association between certain bacteria, which have consistently been linked to CRC (77, 114). While significance levels varied, effect sizes for these taxa, including *F. nucleatum* and *G. morbillorum,* were largely comparable in men and women. These findings suggest that these CRC-associated species appear largely comparable between sexes. Interestingly, *pks*-positive *E. coli* was significantly associated with CRC among women. Most earlier studies have not investigated sex-specific effects of *pks* status on CRC development (114, 115). Our results point to a possible sex-related dimension of *pks* role in CRC that should be further researched.

A major strength of our study is the extensive microbiome dataset, combined with clinically verified outcome data, enabling a thorough assessment of screening-relevant outcomes with minimal risk of misclassification. Additionally, access to detailed lifestyle and demographic data enabled robust covariate adjustments, distinguishing our study from most prior investigations on this topic (108, 109, 116). However, several limitations must be acknowledged. First, the cross-sectional design prevents causal interpretations. Second, participation required enrolment in two consecutive studies: first, the BCSN, followed by CRCbiome, potentially introducing selection bias. In the BCSN, certain discrepancies existed in participant characteristics between men and women. For men, non-participation was linked to younger age and not living with a partner, whereas for women, it was associated with living farther from the screening centre (117, 118). Third, while our comprehensive covariate adjustments strengthen the study, the possibility of residual confounding cannot be ruled out.

The microbial variability observed across lesion subtypes in women, suggests that microbial biomarkers might be particularly effective in women, for whom FIT performance is suggested to be suboptimal. Incorporating microbial markers alongside FIT could reduce false positives and ultimately make screening more equitable across sexes. Our study underscores the importance of validating microbial biomarkers across diverse populations. Beyond CRC, understanding sex-related microbial differences may also have broader implications for other gastrointestinal conditions, including IBD and IBS.

To conclude, in this FIT-positive CRC screening population, our results indicate that sex modifies the association between the microbiome and early stages of colorectal lesions, independent of major lifestyle and demographic factors. This includes marked sex differences in microbial diversity and composition, with 32 bacterial taxa, and 99 functional pathways differing between women and men. Notably, we observed significant microbial differences, including *E. coli pks* status, across lesion subtypes in women, but not in men. Our results underscore the importance of considering sex in research investigating the gut microbiome-colorectal lesion axis, with implications for biomarker discovery and tailoring personalised CRC prevention, detection, and therapy.

## Data Availability

Data from the CRCbiome project have been deposited in the database Federated EGA under accession code EGAS50000000170. Due to the sensitive nature of the data derived from human subjects, including personal health information, analyses and sharing of data from this project must comply with the General Data Protection Regulation (GDPR). How to get access to the data is described here: https://www.mn.uio.no/bils/english/groups/rounge-group/crcbiome/. R-scripts used in this study are available at: https://github.com/Rounge-lab/sex-differences

https://www.ega-archive.org/studies/EGAS50000000170

## Abbreviations

ACME: Average causal mediation effect
ADE: Average direct effect
AICR: American Institute for Cancer Research
ANOVA: Analysis of variance
ASV: Amplicon sequence variant
BCSN: Bowel Cancer Screening in Norway
BH: Benjamini-Hochberg
BMI: Body mass index
CI: Confidence interval
CRC: Colorectal cancer
FFQ: Food frequency questionnaire
FIT: Fecal immunochemical test
GO: Gene ontology
KBS: Kostberegningssystem
MaAsLin: Microbiome multivariable associations with linear models
NCT: National clinical trial
OR: Odds ratio
PCoA: Principal coordinate analysis
PERMANOVA: Permutational multivariate analysis of variance
PPV: Positive predictive value
SD: Standard deviation
TE: Total effect
TSS: Total sum scaling
WCRF: World Cancer Research Fund

## Declarations

### Ethics approval and consent to participate

The BCSN trial and the CRCbiome study have been approved by the Regional Committee for Medical and Health-related Research Ethics in Southeast Norway (REK ref: 2011/1272 and 63148, respectively). All participants provided written informed consent.

## Consent for publication

Not applicable

## Availability of data and materials

The datasets generated and/or analysed during the current study are available in the database Federated EGA under accession code EGAS50000000170. Due to the sensitive nature of the data derived from human subjects, including personal health information, analyses, and sharing of data from this project must comply with the General Data Protection Regulation (GDPR). How to get access to the data is described here: https://www.mn.uio.no/bils/english/groups/rounge-group/crcbiome/. R-scripts used in this study are available at: https://github.com/Rounge-lab/sex-differences

## Author contributions

TBR and PB were responsible for the conception and design of the study. ASK, CBJ, and EB analysed the data. VB performed the sample preparation and lab work. ASK, CBJ, EB, EA, EB, PB, and TBR interpreted the data. ASK and CBJ drafted the manuscript. All authors have revised, commented on, and approved the final submitted manuscript.

## Acknowledgements

We would like to acknowledge Jan Inge Nordby for his contribution to sample preparation and lab work. Library preparation and sequencing were performed at the FIMM Technology Centre supported by HiLIFE and Biocenter Finland. We would especially like to thank Harri A. Kangas, and Pekka J. Ellonen. Finally, we want to thank our project coordinator Maja Sigerseth Jacobsen.

## References

1. Bray F, Laversanne M, Sung H, Ferlay J, Siegel RL, Soerjomataram I, et al. Global cancer statistics 2022: GLOBOCAN estimates of incidence and mortality worldwide for 36 cancers in 185 countries. CA Cancer J Clin. 2024;74(3):229–63.

2. Global Cancer Observatory. Cancer Tomorrow | IARC - https://gco.iarc.who.int/tomorrow [Internet]. [Available from: https://gco.iarc.fr/tomorrow/en/dataviz/trends?types=0_1&sexes=1_2&mode=population&group_populations=0&multiple_populations=1&multiple_cancers=1&cancers=8_41&populations=900&apc=cat_ca20v1.5_ca23v-1.5&group_cancers=1.

3. Sung H, Ferlay J, Siegel RL, Laversanne M, Soerjomataram I, Jemal A, et al. Global Cancer Statistics 2020: GLOBOCAN Estimates of Incidence and Mortality Worldwide for 36 Cancers in 185 Countries. CA Cancer J Clin. 2021;71(3):209–49.

4. Baraibar I, Ros J, Saoudi N, Salva F, Garcia A, Castells MR, et al. Sex and gender perspectives in colorectal cancer. ESMO Open. 2023;8(2):101204.

5. Atkin W, Wooldrage K, Parkin DM, Kralj-Hans I, MacRae E, Shah U, et al. Long term effects of once-only flexible sigmoidoscopy screening after 17 years of follow-up: the UK Flexible Sigmoidoscopy Screening randomised controlled trial. Lancet. 2017;389(10076):1299–311.

6. Schoen RE, Pinsky PF, Weissfeld JL, Yokochi LA, Church T, Laiyemo AO, et al. Colorectal-cancer incidence and mortality with screening flexible sigmoidoscopy. N Engl J Med. 2012;366(25):2345–57.

7. Segnan N, Armaroli P, Bonelli L, Risio M, Sciallero S, Zappa M, et al. Once-only sigmoidoscopy in colorectal cancer screening: follow-up findings of the Italian Randomized Controlled Trial--SCORE. J Natl Cancer Inst. 2011;103(17):1310–22.

8. Holme O, Loberg M, Kalager M, Bretthauer M, Hernan MA, Aas E, et al. Effect of flexible sigmoidoscopy screening on colorectal cancer incidence and mortality: a randomized clinical trial. JAMA. 2014;312(6):606–15.

9. Mandel JS, Bond JH, Church TR, Snover DC, Bradley GM, Schuman LM, et al. Reducing mortality from colorectal cancer by screening for fecal occult blood. Minnesota Colon Cancer Control Study. N Engl J Med. 1993;328(19):1365–71.

10. Kronborg O, Fenger C, Olsen J, Jorgensen OD, Sondergaard O. Randomised study of screening for colorectal cancer with faecal-occult-blood test. Lancet. 1996;348(9040):1467–71.

11. Lindholm E, Brevinge H, Haglind E. Survival benefit in a randomized clinical trial of faecal occult blood screening for colorectal cancer. Br J Surg. 2008;95(8):1029–36.

12. Hardcastle JD, Chamberlain JO, Robinson MH, Moss SM, Amar SS, Balfour TW, et al. Randomised controlled trial of faecal-occult-blood screening for colorectal cancer. Lancet. 1996;348(9040):1472–7.

13. Schreuders EH, Ruco A, Rabeneck L, Schoen RE, Sung JJ, Young GP, et al. Colorectal cancer screening: a global overview of existing programmes. Gut. 2015;64(10):1637–49.

14. Council of the European Union. Council Recommendation on strengthening prevention through early detection: A new EU approach on cancer screening replacing Council Recommendation 2003/878/EC. Off J Eur Union. 2022;100:1–10.

15. Randel KR, Botteri E, de Lange T, Schult AL, Eskeland SL, El-Safadi B, et al. Performance of Faecal Immunochemical Testing for Colorectal Cancer Screening at Varying Positivity Thresholds. Aliment Pharmacol Ther. 2025;61(1):122–31.

16. Ladabaum U, Dominitz JA, Kahi C, Schoen RE. Strategies for Colorectal Cancer Screening. Gastroenterology. 2020;158(2):418–32.

17. Haug U, Kuntz KM, Knudsen AB, Hundt S, Brenner H. Sensitivity of immunochemical faecal occult blood testing for detecting left- vs right-sided colorectal neoplasia. Br J Cancer. 2011;104(11):1779–85.

18. Hultcrantz R. Aspects of Colorectal cancer screening, methods, age and gender. J Intern Med. 2020.

19. Brenner H, Haug U, Hundt S. Sex Differences in Performance of Fecal Occult Blood Testing. 2010;105(11):2457–64.

20. Brenner H, Qian J, Werner S. Variation of diagnostic performance of fecal immunochemical testing for hemoglobin by sex and age: results from a large screening cohort. Clin Epidemiol. 2018;10:381–9.

21. White A, Ironmonger L, Steele RJC, Ormiston-Smith N, Crawford C, Seims A. A review of sex-related differences in colorectal cancer incidence, screening uptake, routes to diagnosis, cancer stage and survival in the UK. BMC cancer. 2018;18(1):906.

22. Njor SH, Rasmussen M, Friis-Hansen L, Andersen B. Varying fecal immunochemical test screening cutoffs by age and gender: a way to increase detection rates and reduce the number of colonoscopies. Gastrointest Endosc. 2022;95(3):540–9.

23. Grobbee EJ, Wieten E, Hansen BE, Stoop EM, de Wijkerslooth TR, Lansdorp-Vogelaar I, et al. Fecal immunochemical test-based colorectal cancer screening: The gender dilemma. United European Gastroenterol J. 2017;5(3):448–54.

24. Kim SE, Paik HY, Yoon H, Lee JE, Kim N, Sung MK. Sex- and gender-specific disparities in colorectal cancer risk. World J Gastroenterol. 2015;21(17):5167–75.

25. Streett SE. Endoscopic colorectal cancer screening in women: can we do better? Gastrointest Endosc. 2007;65(7):1047–9.

26. Witte TN, Enns R. The difficult colonoscopy. Can J Gastroenterol. 2007;21(8):487–90.

27. Holme O, Loberg M, Kalager M, Bretthauer M, Hernan MA, Aas E, et al. Long-Term Effectiveness of Sigmoidoscopy Screening on Colorectal Cancer Incidence and Mortality in Women and Men: A Randomized Trial. Ann Intern Med. 2018;168(11):775–82.

28. Schwabe RF, Jobin C. The microbiome and cancer. Nat Rev Cancer. 2013;13(11):800–12.

29. DeWeerdt S. Microbiome: Microbial mystery. Nature. 2015;521(7551):S10–1.

30. Garrett WS. The gut microbiota and colon cancer. Science. 2019;364(6446):1133–5.

31. Wong CC, Yu J. Gut microbiota in colorectal cancer development and therapy. Nat Rev Clin Oncol. 2023;20(7):429–52.

32. Wong SH, Yu J. Gut microbiota in colorectal cancer: mechanisms of action and clinical applications. Nat Rev Gastroenterol Hepatol. 2019;16(11):690–704.

33. Janney A, Powrie F, Mann EH. Host-microbiota maladaptation in colorectal cancer. Nature. 2020;585(7826):509–17.

34. Xing C, Du Y, Duan T, Nim K, Chu J, Wang HY, et al. Interaction between microbiota and immunity and its implication in colorectal cancer. Front Immunol. 2022;13:963819.

35. Chen Y, Chen YX. Microbiota-Associated Metabolites and Related Immunoregulation in Colorectal Cancer. Cancers (Basel). 2021;13(16):4054.

36. Zheng DP, Liwinski T, Elinav E. Interaction between microbiota and immunity in health and disease. Cell Res. 2020;30(6):492–506.

37. Park CH, Eun CS, Han DS. Intestinal microbiota, chronic inflammation, and colorectal cancer. Intest Res. 2018;16(3):338–45.

38. Parker BJ, Wearsch PA, Veloo ACM, Rodriguez-Palacios A. The Genus Alistipes: Gut Bacteria With Emerging Implications to Inflammation, Cancer, and Mental Health. Front Immunol. 2020;11:906.

39. Sanchez-Alcoholado L, Ordonez R, Otero A, Plaza-Andrade I, Laborda-Illanes A, Medina JA, et al. Gut Microbiota-Mediated Inflammation and Gut Permeability in Patients with Obesity and Colorectal Cancer. Int J Mol Sci. 2020;21(18).

40. Wu Z, Huang Y, Zhang R, Zheng C, You F, Wang M, et al. Sex differences in colorectal cancer: with a focus on sex hormone-gut microbiome axis. Cell Commun Signal. 2024;22(1):167.

41. Rodriguez-Santiago Y, Garay-Canales CA, Nava-Castro KE, Morales-Montor J. Sexual dimorphism in colorectal cancer: molecular mechanisms and treatment strategies. Biol Sex Differ. 2024;15(1):48.

42. Zhou YL, Hong J. Gut microbiota: Guardians of the female gut health. Cancer Cell. 2023;41(8):1392–4.

43. Cheng WT, Kantilal HK, Davamani F. The Mechanism of Bacteroides fragilis Toxin Contributes to Colon Cancer Formation. Malays J Med Sci. 2020;27(4):9–21.

44. Spigaglia P, Barbanti F, Germinario EAP, Criscuolo EM, Bruno G, Sanchez-Mete L, et al. Comparison of microbiological profile of enterotoxigenic Bacteroides fragilis (ETBF) isolates from subjects with colorectal cancer (CRC) or intestinal pre-cancerous lesions versus healthy individuals and evaluation of environmental factors involved in intestinal dysbiosis. Anaerobe. 2023;82:102757.

45. Arima K, Zhong R, Ugai T, Zhao M, Haruki K, Akimoto N, et al. Western-Style Diet, pks Island-Carrying Escherichia coli, and Colorectal Cancer: Analyses From Two Large Prospective Cohort Studies. Gastroenterology. 2022;163(4):862–74.

46. Pleguezuelos-Manzano C, Puschhof J, Rosendahl Huber A, van Hoeck A, Wood HM, Nomburg J, et al. Mutational signature in colorectal cancer caused by genotoxic pks(+) E. coli. Nature. 2020;580(7802):269–73.

47. Sengupta S, Muir JG, Gibson PR. Does butyrate protect from colorectal cancer? J Gastroenterol Hepatol. 2006;21(1 Pt 2):209–18.

48. Wirbel J, Pyl PT, Kartal E, Zych K, Kashani A, Milanese A, et al. Meta-analysis of fecal metagenomes reveals global microbial signatures that are specific for colorectal cancer. Nat Med. 2019;25(4):679–89.

49. Chen D, Jin D, Huang S, Wu J, Xu M, Liu T, et al. Clostridium butyricum, a butyrate-producing probiotic, inhibits intestinal tumor development through modulating Wnt signaling and gut microbiota. Cancer Lett. 2020;469:456–67.

50. Zwezerijnen-Jiwa FH, Sivov H, Paizs P, Zafeiropoulou K, Kinross J. A systematic review of microbiome-derived biomarkers for early colorectal cancer detection. Neoplasia. 2023;36:100868.

51. Fusco W, Bricca L, Kaitsas F, Tartaglia MF, Venturini I, Rugge M, et al. Gut microbiota in colorectal cancer: From pathogenesis to clinic. Best Pract Res Clin Gastroenterol. 2024;72:101941.

52. Birkeland EE, Kvaerner AS, Avershina E, Bucher-Johannessen C, Bemanian V, Blix HS, et al. Microbiome signatures for detection of colorectal lesions in population-based FIT screening. medRxiv [Preprint]. 2025.

53. Valeri F, Endres K. How biological sex of the host shapes its gut microbiota. Front Neuroendocrinol. 2021;61:100912.

54. Vemuri R, Sylvia KE, Klein SL, Forster SC, Plebanski M, Eri R, et al. The microgenderome revealed: sex differences in bidirectional interactions between the microbiota, hormones, immunity and disease susceptibility. Semin Immunopathol. 2019;41(2):265–75.

55. Yoon K, Kim N. Roles of Sex Hormones and Gender in the Gut Microbiota. J Neurogastroenterol Motil. 2021;27(3):314–25.

56. Thackray VG. Sex, Microbes, and Polycystic Ovary Syndrome. Trends Endocrinol Metab. 2019;30(1):54–65.

57. Kvaerner AS, Birkeland E, Bucher-Johannessen C, Vinberg E, Nordby JI, Kangas H, et al. The CRCbiome study: a large prospective cohort study examining the role of lifestyle and the gut microbiome in colorectal cancer screening participants. BMC cancer. 2020.

58. Randel KR, Schult AL, Botteri E, Hoff G, Bretthauer M, Ursin G, et al. Colorectal Cancer Screening With Repeated Fecal Immunochemical Test Versus Sigmoidoscopy: Baseline Results From a Randomized Trial. Gastroenterology. 2021;160(4):1085–96 e5.

59. Andersen LF, Solvoll K, Johansson LR, Salminen I, Aro A, Drevon CA. Evaluation of a food frequency questionnaire with weighed records, fatty acids, and alpha-tocopherol in adipose tissue and serum. American journal of epidemiology. 1999;150(1):75–87.

60. Andersen LF, Tomten H, Haggarty P, Lovo A, Hustvedt BE. Validation of energy intake estimated from a food frequency questionnaire: a doubly labelled water study. Eur J Clin Nutr. 2003;57(2):279–84.

61. Andersen LF, Veierod MB, Johansson L, Sakhi A, Solvoll K, Drevon CA. Evaluation of three dietary assessment methods and serum biomarkers as measures of fruit and vegetable intake, using the method of triads. Br J Nutr. 2005;93(4):519–27.

62. Carlsen MH, Lillegaard IT, Karlsen A, Blomhoff R, Drevon CA, Andersen LF. Evaluation of energy and dietary intake estimates from a food frequency questionnaire using independent energy expenditure measurement and weighed food records. Nutr J. 2010;9(1):37.

63. Brunvoll SH, Thune I, Frydenberg H, Flote VG, Bertheussen GF, Schlichting E, et al. Validation of repeated self-reported n-3 PUFA intake using serum phospholipid fatty acids as a biomarker in breast cancer patients during treatment. Nutr J. 2018;17(1):94.

64. Nes M, Frost Andersen L, Solvoll K, Sandstad B, Hustvedt BE, Lovo A, et al. Accuracy of a quantitative food frequency questionnaire applied in elderly Norwegian women. Eur J Clin Nutr. 1992;46(11):809–21.

65. Norwegian Food Safety Authority. Norwegian Food Composition Database 2019 [Internet]. 2019 [cited 2025 20.02]. Available from: www.matvaretabellen.no.

66. Shams-White MM, Brockton NT, Mitrou P, Romaguera D, Brown S, Bender A, et al. Operationalizing the 2018 World Cancer Research Fund/American Institute for Cancer Research (WCRF/AICR) Cancer Prevention Recommendations: A Standardized Scoring System. Nutrients. 2019;11(7).

67. Shams-White MM, Romaguera D, Mitrou P, Reedy J, Bender A, Brockton NT. Further Guidance in Implementing the Standardized 2018 World Cancer Research Fund/American Institute for Cancer Research (WCRF/AICR) Score. Cancer Epidemiology Biomarkers & Prevention. 2020;29(5):889–94.

68. Kvaerner AS, Andersen AR, Henriksen HB, Knudsen MD, Johansen AMW, Hjartaker A, et al. Associations of the 2018 World Cancer Research Fund/American Institute of Cancer Research (WCRF/AICR) cancer prevention recommendations with stages of colorectal carcinogenesis. Cancer Med. 2023;12(13):14806–19.

69. WHO. Collaborating centre for drug statistics methodology. ATC index with DDDs. Oslo: WHO. 1997.

70. Istvan P, Birkeland E, Avershina E, Kvaerner AS, Bemanian V, de Vos W, et al. Exploring the gut virome in fecal immunochemical test stool samples reveals novel associations with lifestyle in a large population-based study. medRxiv. 2023:2023.08. 24.23294548.

71. Rounge TB, Meisal R, Nordby JI, Ambur OH, de Lange T, Hoff G. Evaluating gut microbiota profiles from archived fecal samples. BMC Gastroenterol. 2018;18(1):171.

72. Blanco-Miguez A, Beghini F, Cumbo F, McIver LJ, Thompson KN, Zolfo M, et al. Extending and improving metagenomic taxonomic profiling with uncharacterized species using MetaPhlAn 4. Nat Biotechnol. 2023;41(11):1633–44.

73. Beghini F, McIver LJ, Blanco-Miguez A, Dubois L, Asnicar F, Maharjan S, et al. Integrating taxonomic, functional, and strain-level profiling of diverse microbial communities with bioBakery 3. Elife. 2021;10.

74. Wang Y, Wang Q, Huang H, Huang W, Chen Y, McGarvey PB, et al. A crowdsourcing open platform for literature curation in UniProt. PLoS Biol. 2021;19(12):e3001464.

75. Ashburner M, Ball CA, Blake JA, Botstein D, Butler H, Cherry JM, et al. Gene ontology: tool for the unification of biology. The Gene Ontology Consortium. Nat Genet. 2000;25(1):25–9.

76. Aleksander SA, Balhoff J, Carbon S, Cherry JM, Drabkin HJ, Ebert D, et al. Gene Ontology Consortium. The Gene Ontology knowledgebase in 2023. Genetics. 2023;224(1).

77. Piccinno G, Thompson KN, Manghi P, Ghazi AR, Thomas AM, Blanco-Miguez A, et al. Pooled analysis of 3,741 stool metagenomes from 18 cohorts for cross-stage and strain-level reproducible microbial biomarkers of colorectal cancer. Nat Med. 2025;31(7):2416–29.

78. Van Buuren S, Groothuis-Oudshoorn K. mice: Multivariate imputation by chained equations in R. Journal of statistical software. 2011;45:1–67.

79. Oksanen J, Blanchet FG, Kindt R, Legendre P, Minchin P, O’Hara R, et al. Vegan: community ecology package. Ordination methods, diversity analysis and other functions for community and vegetation ecologists. 2015:2.3–1.

80. Mallick H, Rahnavard A, McIver LJ, Ma S, Zhang Y, Nguyen LH, et al. Multivariable association discovery in population-scale meta-omics studies. PLoS Comput Biol. 2021;17(11):e1009442.

81. Wickham H, Averick M, Bryan J, Chang W, McGowan LDA, François R, et al. Welcome to the Tidyverse. Journal of open source software. 2019;4(43):1686.

82. Kassambara A. rstatix: Pipe-friendly framework for basic statistical tests. R package version 06 0. 2020.

83. Venables WN, Ripley BD. Modern applied statistics with S-PLUS: Springer Science & Business Media; 2013.

84. Thomas AM, Manghi P, Asnicar F, Pasolli E, Armanini F, Zolfo M, et al. Metagenomic analysis of colorectal cancer datasets identifies cross-cohort microbial diagnostic signatures and a link with choline degradation. Nat Med. 2019;25(4):667–78.

85. Young C, Wood HM, Fuentes Balaguer A, Bottomley D, Gallop N, Wilkinson L, et al. Microbiome Analysis of More Than 2,000 NHS Bowel Cancer Screening Programme Samples Shows the Potential to Improve Screening Accuracy. Clin Cancer Res. 2021;27(8):2246–54.

86. Yachida S, Mizutani S, Shiroma H, Shiba S, Nakajima T, Sakamoto T, et al. Metagenomic and metabolomic analyses reveal distinct stage-specific phenotypes of the gut microbiota in colorectal cancer. Nat Med. 2019;25(6):968–76.

87. Botteri E, Iodice S, Bagnardi V, Raimondi S, Lowenfels AB, Maisonneuve P. Smoking and colorectal cancer: a meta-analysis. JAMA. 2008;300(23):2765–78.

88. Bailie L, Loughrey MB, Coleman HG. Lifestyle risk factors for serrated colorectal polyps: a systematic review and meta-analysis. Gastroenterology. 2017;152(1):92–104.

89. Clinton SK, Giovannucci EL, Hursting SD. The world cancer research fund/American institute for cancer research third expert report on diet, nutrition, physical activity, and cancer: impact and future directions. The Journal of nutrition. 2020;150(4):663–71.

90. Kvaerner AS, Birkeland E, Vinberg E, Hoff G, Hjartaker A, Rounge TB, et al. Associations of red and processed meat intake with screen-detected colorectal lesions. Br J Nutr. 2023;129(12):2122–32.

91. Kvaerner AS, Birkeland E, Avershina E, Botteri E, Bucher-Johannessen C, Dines Knudsen M, et al. The association between alcohol consumption and colorectal carcinogenesis is partially mediated by the gut microbiome. medRxiv [Preprint]. 2024:2024.10. 17.24315656.

92. Knudsen MD, Kvaerner AS, Botteri E, Holme O, Hjartaker A, Song M, et al. Lifestyle predictors for inconsistent participation to fecal based colorectal cancer screening. BMC cancer. 2022;22(1):172.

93. Knudsen MD, Botteri E, Holme O, Hjartaker A, Song M, Thiis-Evensen E, et al. Association between lifestyle and site-specific advanced colorectal lesions in screening with faecal immunochemical test and sigmoidoscopy. Dig Liver Dis. 2021;53(3):353–9.

94. Joo HJ, Lee HS, Jang BI, Kim DB, Kim JH, Park JJ, et al. Sex-specific differences in colorectal cancer: A multicenter retrospective cohort study. Cancer Reports. 2023;6(8):e1845.

95. Niedermaier T, Heisser T, Gies A, Guo F, Amitay EL, Hoffmeister M, et al. To what extent is male excess risk of advanced colorectal neoplasms explained by known risk factors? Results from a large German screening population. Intl Journal of Cancer. 2021;149(11):1877–86.

96. Selby K, Levine EH, Doan C, Gies A, Brenner H, Quesenberry C, et al. Effect of Sex, Age, and Positivity Threshold on Fecal Immunochemical Test Accuracy: A Systematic Review and Meta-analysis. Gastroenterology. 2019;157(6):1494–505.

97. Zhang X, Zhong H, Li Y, Shi Z, Ren H, Zhang Z, et al. Sex- and age-related trajectories of the adult human gut microbiota shared across populations of different ethnicities. Nat Aging. 2021;1(1):87–100.

98. Li Q, Chan H, Liu WX, Liu CA, Zhou Y, Huang D, et al. Carnobacterium maltaromaticum boosts intestinal vitamin D production to suppress colorectal cancer in female mice. Cancer Cell. 2023;41(8):1450–65 e8.

99. Song CH, Kim N, Nam RH, Choi SI, Lee HN, Surh YJ. 17beta-Estradiol supplementation changes gut microbiota diversity in intact and colorectal cancer-induced ICR male mice. Scientific reports. 2020;10(1):12283.

100. Bucher-Johannessen C, Senthakumaran T, Avershina E, Birkeland E, Hoff G, Bemanian V, et al. Species-level verification of Phascolarctobacterium association with colorectal cancer. mSystems. 2024;9(10):e0073424.

101. Bucher-Johannessen C, Birkeland EE, Vinberg E, Bemanian V, Hoff G, Berstad P, et al. Long-term follow-up of colorectal cancer screening attendees identifies differences in Phascolarctobacterium spp. using 16S rRNA and metagenome sequencing. Front Oncol. 2023;13:1183039.

102. Senthakumaran T, Moen AEF, Tannaes TM, Endres A, Brackmann SA, Rounge TB, et al. Microbial dynamics with CRC progression: a study of the mucosal microbiota at multiple sites in cancers, adenomatous polyps, and healthy controls. Eur J Clin Microbiol Infect Dis. 2023;42(3):305–22.

103. Hua H, Sun Y, He X, Chen Y, Teng L, Lu C. Intestinal Microbiota in Colorectal Adenoma-Carcinoma Sequence. Front Med (Lausanne). 2022;9:888340.

104. Doden HL, Pollet RM, Mythen SM, Wawrzak Z, Devendran S, Cann I, et al. Structural and biochemical characterization of 20beta-hydroxysteroid dehydrogenase from Bifidobacterium adolescentis strain L2-32. The Journal of biological chemistry. 2019;294(32):12040–53.

105. Sinha T, Vich Vila A, Garmaeva S, Jankipersadsing SA, Imhann F, Collij V, et al. Analysis of 1135 gut metagenomes identifies sex-specific resistome profiles. Gut Microbes. 2019;10(3):358–66.

106. McCurry MD, D’Agostino GD, Walsh JT, Bisanz JE, Zalosnik I, Dong X, et al. Gut bacteria convert glucocorticoids into progestins in the presence of hydrogen gas. Cell. 2024;187(12):2952–68 e13.

107. Jiang L, Fei H, Tong J, Zhou J, Zhu J, Jin X, et al. Hormone Replacement Therapy Reverses Gut Microbiome and Serum Metabolome Alterations in Premature Ovarian Insufficiency. Front Endocrinol (Lausanne). 2021;12:794496.

108. Liao H, Li C, Ai Y, Kou Y. Gut microbiome is more stable in males than in females during the development of colorectal cancer. J Appl Microbiol. 2021;131(1):435–48.

109. Yang X, Li P, Qu Z, Zhuang J, Wu Y, Wu W, et al. Gut bacteria and sex differences in colorectal cancer. J Med Microbiol. 2023;72(6).

110. Song CH, Kim N, Nam RH, Choi SI, Jang JY, Kim EH, et al. The Possible Preventative Role of Lactate- and Butyrate-Producing Bacteria in Colorectal Carcinogenesis. Gut Liver. 2024;18(4):654–66.

111. Yu S, Chu J, Wu Y, Zhuang J, Qu Z, Song Y, et al. Third-generation PacBio sequencing to explore gut bacteria and gender in colorectal cancer. Microb Pathog. 2024;192:106684.

112. Wang SY, Dong XT, Yuan Z, Jin LX, Gao WF, Han YK, et al. Factors associated with false fecal immunochemical test results in colorectal cancer screening. World J Gastrointest Oncol. 2025;17(4):101487.

113. Whang A, Nagpal R, Yadav H. Bi-directional drug-microbiome interactions of anti-diabetics. EBioMedicine. 2019;39:591–602.

114. Gaab ME, Lozano PO, Ibanez D, Manese KD, Riego FM, Tiongco RE, et al. A Meta-Analysis on the Association of Colibactin-Producing pks+ Escherichia coli with the Development of Colorectal Cancer. Lab Med. 2023;54(1):75–82.

115. de Klaver W, de Wit M, Bolijn A, Tijssen M, Delis-van Diemen P, Lemmens M, et al. Polyketide synthase positive Escherichia coli one-time measurement in stool is not informative of colorectal cancer risk in a screening setting. J Pathol. 2024;263(2):217–25.

116. Lattanzi G, Perillo F, Diaz-Basabe A, Caridi B, Amoroso C, Baeri A, et al. Estrogen-related differences in antitumor immunity and gut microbiome contribute to sexual dimorphism of colorectal cancer. Oncoimmunology. 2024;13(1):2425125.

117. Berthelsen M, Berstad P, Randel KR, Hoff G, Natvig E, Holme O, et al. The impact of driving time on participation in colorectal cancer screening with sigmoidoscopy and faecal immunochemical blood test. Cancer Epidemiol. 2022;80:102244.

118. Botteri E, Hoff G, Randel KR, Holme O, de Lange T, Bernklev T, et al. Characteristics of nonparticipants in a randomised colorectal cancer screening trial comparing sigmoidoscopy and faecal immunochemical testing. Int J Cancer. 2022;151(3):361–71.

