## Supplementary materials for "Gut microbiome signatures of colorectal cancer development are more pronounced in women compared to men in a population-based screening cohort"

**Supplementary material: Women and men exhibit distinct gut microbial profiles linked to colorectal cancer development**

*Bucher-Johannessen, C. [1,2,3]*, Kværner, AS.[4]*, Birkeland, E.[5], Botteri E.[2,4], Avershina, E.[1,5], Bemanian, V.[6], Hoff, G.[4,7], Randel, KR.[4], Hovig, E.[1,3], Berstad, P.[4], Rounge, TB.[1,4,5]*

**Shared first authorship*

Supplementary Data 1: Differentially abundant species and functions between men and women.

Supplementary Data 2: Differentially abundant species and functions between colonoscopy findings, stratified on sex.

**Supplementary Table 1:** Participant characteristics in the control group.

| Characteristic[1] | N | Women (N = 219) | Men (N = 222) | Overall (N = 441) | p-value[2] |
|---|---|---|---|---|---|
| Age years | 441 | 66.2 (60.7, 71.0) | 65.8 (60.9, 71.6) | 65.9 (60.8, 71.1) | 0.891 |
| Screening center, n (%) | 441 | | | | 0.367 |
| Moss | | 136 (62%) | 147 (66%) | 283 (64%) | |
| Bærum | | 83 (38%) | 75 (34%) | 158 (36%) | |
| Nationality, n (%) | 427 | | | | 0.567 |
| Native | | 199 (93%) | 193 (91%) | 392 (92%) | |
| Non-native | | 16 (7.4%) | 19 (9.0%) | 35 (8.2%) | |
| Family history of CRC, n (%) | 391 | 41 (21%) | 28 (15%) | 69 (18%) | 0.119 |
| Marital status, n (%) | 435 | | | | **0.001** |
| Not married/non-cohabiting | | 48 (22%) | 23 (11%) | 71 (16%) | |
| Married/cohabiting | | 169 (78%) | 195 (89%) | 364 (84%) | |
| Education, n (%) | 431 | | | | 0.223 |
| Primary school | | 52 (24%) | 38 (18%) | 90 (21%) | |
| High school | | 77 (36%) | 88 (41%) | 165 (38%) | |
| University/college | | 86 (40%) | 90 (42%) | 176 (41%) | |
| Employment status, n (%) | 436 | | | | **0.011** |
| Employed | | 64 (29%) | 90 (41%) | 154 (35%) | |
| Retired | | 108 (50%) | 102 (47%) | 210 (48%) | |
| Other | | 45 (21%) | 27 (12%) | 72 (17%) | |
| Bowel disorder, n (%) | 427 | | | | **0.012** |
| No bowel disease | | 163 (77%) | 187 (87%) | 350 (82%) | |
| Any disorder | | 48 (23%) | 29 (13%) | 77 (18%) | |
| colo_IBD, n (%) | 441 | 9 (4.1%) | 11 (5.0%) | 20 (4.5%) | 0.67 |
| Smoking status, n (%) | 434 | | | | 0.114 |
| Never smoked | | 91 (42%) | 75 (35%) | 166 (38%) | |
| Ever smoked | | 126 (58%) | 142 (65%) | 268 (62%) | |
| Snus, n (%)[3] | 419 | | | | **<0.001** |
| Non snuser | | 206 (99%) | 184 (87%) | 390 (93%) | |
| Snuser | | 2 (1.0%) | 27 (13%) | 29 (6.9%) | |
| Energy , Median (IQR) | 428 | 1,892 (1,556, 2,389) | 2,418 (1,951, 2,835) | 2,154 (1,705, 2,653) | **<0.001** |
| BMI, Median (IQR) | 426 | 25.5 (22.9, 28.9) | 26.9 (24.2, 28.7) | 26.3 (23.7, 28.7) | **0.004** |
| Physical activity min/week | 436 | 105 (0, 300) | 180 (0, 390) | 135 (0, 368) | 0.098 |
| Antibiotic use last 3 months, n (%)[4] | 411 | 35 (17%) | 23 (11%) | 58 (14%) | 0.093 |
| FIT value, Median (IQR) | 441 | 163 (104, 356) | 175 (108, 304) | 166 (106, 326) | 0.752 |
| WCRF/AICR score, Median (IQR)[5] | 405 | 4.00 (3.00, 4.50) | 3.50 (2.75, 4.25) | 3.75 (3.00, 4.50) | **0.001** |
| reads_proc, Median (IQR) | 441 | 12,546,270 (10,527,614, 14,458,304) | 12,844,464 (11,033,280, 14,802,455) | 12,684,129 (10,725,253, 14,638,307) | 0.172 |

*[1]Values are median (IQR) for continuous variables and n (%) for categorical variables.*
*[2]P-values are derived from Wilcoxon rank-sum test, Students t-testPearson`s Chi-squared testing, as appropriate.*
*[3]To be defined as a snuser one had to be a regular or occasional user or having quit consumption within the last ten years.*
*[4]Recent antibiotic use was defined based on self-reported use of antibiotics within 3 months of sample collection.*

*[5]WCRF/AICR score was a seven point score that measured adherence to the World Cancer Research Fund/American Institute of Cancer Research (WCRF/AICR) cancer prevention recommendations.*
*Abbreviations: AICR; American Institute of Cancer Research, BMI; Body mass index, CRC; colorectal cancer, g; gram, n; number of participants in a group, WCRF; World Cancer Research Fund*

**Supplementary Table 2:** Participant characteristics for women and men separately by screening colonoscopy findings[1].

| | | Women | | | | | | Men | | | | |
|---|---|---|---|---|---|---|---|---|---|---|---|---|
| **Characteristic[1]** | **N** | **No adenoma (N = 221)** | **Non-advanced adenoma (N = 55)** | **Advanced lesion (N = 176)** | **Overall (N = 452)** | **p-value[2]** | **N** | **No adenoma (N = 229)** | **Non-advanced adenoma (N = 87)** | **Advanced lesion (N = 266)** | **Overall (N = 582)** | **p-value[2]** |
| Age, years | 452 | 66.2 (60.8, 71.1) | 69.5 (65.0, 71.4) | 67.8 (62.5, 73.0) | 67.3 (61.8, 71.7) | **0.032** | 582 | 65.8 (60.8, 71.7) | 70.6 (64.5, 74.0) | 67.6 (62.5, 71.7) | 67.0 (61.9, 72.1) | **0.003** |
| Screening center, n (%) | 452 | | | | | **0.02** | 582 | | | | | **<0.001** |
| Moss | | 137 (62%) | 27 (49%) | 86 (49%) | 250 (55%) | | | 149 (65%) | 40 (46%) | 129 (48%) | 318 (55%) | |
| Bærum | | 84 (38%) | 28 (51%) | 90 (51%) | 202 (45%) | | | 80 (35%) | 47 (54%) | 137 (52%) | 264 (45%) | |
| Nationality, n (%) | 442 | | | | | 0.255 | 547 | | | | | 0.191 |
| Native | | 200 (92%) | 53 (98%) | 162 (95%) | 415 (94%) | | | 199 (91%) | 72 (89%) | 233 (94%) | 504 (92%) | |
| Non-native | | 17 (7.8%) | 1 (1.9%) | 9 (5.3%) | 27 (6.1%) | | | 20 (9.1%) | 9 (11%) | 14 (5.7%) | 43 (7.9%) | |
| Family history of CRC, n (%) | 414 | 41 (20%) | 11 (22%) | 35 (21%) | 87 (21%) | 0.953 | 509 | 29 (15%) | 18 (24%) | 51 (22%) | 98 (19%) | 0.11 |
| Localization, n (%) | 452 | | | | | **<0.001** | 582 | | | | | **<0.001** |
| Both | | 0 (0%) | 35 (64%) | 66 (38%) | 101 (22%) | | | 0 (0%) | 57 (66%) | 127 (48%) | 184 (32%) | |
| Distal | | 0 (0%) | 4 (7.3%) | 57 (32%) | 61 (13%) | | | 0 (0%) | 5 (5.7%) | 84 (32%) | 89 (15%) | |
| Negative | | 221 (100%) | 0 (0%) | 0 (0%) | 221 (49%) | | | 229 (100%) | 0 (0%) | 0 (0%) | 229 (39%) | |
| Proximal | | 0 (0%) | 16 (29%) | 53 (30%) | 69 (15%) | | | 0 (0%) | 25 (29%) | 55 (21%) | 80 (14%) | |
| Advanced serrated lesions, n (%) | 452 | | | | | **<0.001** | 582 | | | | | **<0.001** |
| No | | 221 (100%) | 55 (100%) | 116 (66%) | 392 (87%) | | | 229 (100%) | 87 (100%) | 202 (76%) | 518 (89%) | |
| Yes | | 0 (0%) | 0 (0%) | 60 (34%) | 60 (13%) | | | 0 (0%) | 0 (0%) | 64 (24%) | 64 (11%) | |
| Marital status, n (%) | 447 | | | | | **0.047** | 566 | | | | | 0.064 |
| Not married/non-cohabiting | | 48 (22%) | 19 (35%) | 54 (31%) | 121 (27%) | | | 23 (10%) | 13 (15%) | 45 (18%) | 81 (14%) | |
| Married/cohabiting | | 171 (78%) | 35 (65%) | 120 (69%) | 326 (73%) | | | 202 (90%) | 74 (85%) | 209 (82%) | 485 (86%) | |
| Education, n (%) | 444 | | | | | 0.273 | 564 | | | | | 0.848 |
| Primary school | | 52 (24%) | 16 (30%) | 34 (20%) | 102 (23%) | | | 39 (17%) | 14 (16%) | 41 (16%) | 94 (17%) | |
| High school | | 77 (35%) | 21 (40%) | 76 (44%) | 174 (39%) | | | 91 (41%) | 31 (36%) | 97 (38%) | 219 (39%) | |
| University/college | | 88 (41%) | 16 (30%) | 64 (37%) | 168 (38%) | | | 93 (42%) | 42 (48%) | 116 (46%) | 251 (45%) | |
| Employment status, n (%) | 447 | | | | | **0.003** | 568 | | | | | **0.047** |
| Employed | | 64 (29%) | 8 (15%) | 47 (27%) | 119 (27%) | | | 93 (41%) | 25 (29%) | 85 (33%) | 203 (36%) | |
| Retired | | 110 (50%) | 39 (72%) | 110 (63%) | 259 (58%) | | | 105 (46%) | 55 (63%) | 147 (58%) | 307 (54%) | |
| Other | | 45 (21%) | 7 (13%) | 17 (9.8%) | 69 (15%) | | | 28 (12%) | 7 (8.0%) | 23 (9.0%) | 58 (10%) | |
| Bowel disorder, n (%) | 438 | | | | | 0.105 | 564 | | | | | 0.528 |
| No bowel disease | | 165 (77%) | 45 (85%) | 147 (85%) | 357 (82%) | | | 194 (87%) | 77 (90%) | 230 (90%) | 501 (89%) | |
| Any disorder | | 48 (23%) | 8 (15%) | 25 (15%) | 81 (18%) | | | 29 (13%) | 9 (10%) | 25 (9.8%) | 63 (11%) | |
| Smoking status, n (%) | 447 | | | | | 0.371 | 566 | | | | | 0.528 |
| Never smoked | | 92 (42%) | 19 (35%) | 62 (36%) | 173 (39%) | | | 79 (35%) | 36 (41%) | 89 (35%) | 204 (36%) | |
| Ever smoked | | 127 (58%) | 35 (65%) | 112 (64%) | 274 (61%) | | | 145 (65%) | 51 (59%) | 166 (65%) | 362 (64%) | |

| | | | | | | | | | | | | |
|---|---|---|---|---|---|---|---|---|---|---|---|---|
| Snus, n (%)[3] | 418 | | | | | 0.259 | 553 | | | | | 0.386 |
| Non-snuser | | 208 (99%) | 48 (96%) | 156 (99%) | 412 (99%) | | | 190 (87%) | 77 (93%) | 222 (88%) | 489 (88%) | |
| Snuser | | 2 (1.0%) | 2 (4.0%) | 2 (1.3%) | 6 (1.4%) | | | 28 (13%) | 6 (7.2%) | 30 (12%) | 64 (12%) | |
| Energy | 441 | 1,889 (1,553, 2,387) | 1,838 (1,449, 2,190) | 1,998 (1,666, 2,394) | 1,914 (1,572, 2,374) | 0.135 | 566 | 2,405 (1,942, 2,836) | 2,293 (1,852, 2,795) | 2,452 (1,953, 2,949) | 2,399 (1,912, 2,900) | 0.15 |
| Alcohol | 441 | 4 (0, 11) | 5 (0, 13) | 7 (2, 16) | 5 (1, 12) | **0.002** | 566 | 11 (3, 21) | 12 (2, 28) | 14 (6, 26) | 12 (4, 24) | **0.031** |
| BMI | 438 | 25.5 (22.9, 28.7) | 27.5 (23.9, 30.1) | 25.9 (23.4, 28.3) | 25.9 (23.1, 28.8) | 0.097 | 562 | 26.9 (24.2, 28.7) | 27.5 (25.2, 29.4) | 27.1 (24.8, 29.4) | 27.1 (24.6, 29.3) | 0.161 |
| Physical activity min/week | 447 | 135 (0, 300) | 105 (0, 300) | 135 (0, 300) | 135 (0, 300) | 0.444 | 568 | 180 (4, 390) | 105 (8, 300) | 105 (8, 308) | 105 (0, 360) | 0.064 |
| Antibiotic use last 3 months, n (%)[4] | 426 | 36 (17%) | 7 (13%) | 26 (16%) | 69 (16%) | 0.775 | 538 | 24 (11%) | 9 (11%) | 34 (14%) | 67 (12%) | 0.636 |
| FIT value | 452 | 163 (104, 353) | 156 (112, 218) | 221 (147, 571) | 178 (118, 384) | **<0.001** | 582 | 168 (108, 303) | 161 (111, 342) | 206 (127, 592) | 192 (115, 410) | **0.002** |
| WCRF/AICR score | 417 | 4.00 (3.00, 4.50) | 3.50 (2.81, 4.25) | 3.75 (3.00, 4.50) | 3.75 (3.00, 4.50) | 0.065 | 528 | 3.50 (2.75, 4.25) | 3.25 (2.50, 4.00) | 3.50 (2.50, 4.00) | 3.50 (2.75, 4.00) | 0.189 |
| Sequencing depth | 452 | 12,618,983 (10,546,013, 14,572,873) | 11,709,612 (9,737,805, 13,825,396) | 12,789,724 (10,851,340, 14,985,666) | 12,656,840 (10,524,188, 14,576,042) | **0.042** | 582 | 12,788,165 (11,089,612, 14,718,880) | 12,850,996 (10,770,138, 14,642,185) | 12,362,380 (10,665,298, 14,421,430) | 12,599,426 (10,830,200, 14,566,860) | 0.344 |

*[1]Values are median (IQR) for continuous variables and n (%) for categorical variables.*

*[2]P-values are derived from Wilcoxon rank-sum test, Students t-testPearson`s Chi-squared testing, as appropriate.*

*[3]To be defined as a snuser one had to be a regular or occasional user or having quit consumption within the last ten years.*

*[4]Recent antibiotic use was defined based on self-reported use of antibiotics within 3 months of sample collection.*

*[5]WCRF/AICR score was a seven point score that measured adherence to the World Cancer Research Fund/American Institute of Cancer Research (WCRF/AICR) cancer prevention recommendations.*

*Abbreviations: AICR; American Institute of Cancer Research, BMI; Body mass index, CRC; colorectal cancer, g; gram, n; number of participants in a group, WCRF; World Cancer Research Fund*

**Supplementary Table 3**: Showing GI-related symptoms recorded by endoscopist at time of colonoscopy examination and prescription drug use with indication of common comorbidities across lesion subtypes in women and men separately by screening colonoscopy findings.

| | Women | | | | | | Men | | | | | |
|---|---|---|---|---|---|---|---|---|---|---|---|---|
| **Characteristic** | **N** | **No adenoma (N = 221)** | **Non-advanced adenoma (N = 55)** | **Advanced lesion (N = 176)** | **Overall (N = 452)** | **p-value[3]** | **N** | **No adenoma (N = 221)** | **Non-advanced adenoma (N = 55)** | **Advanced lesion (N = 176)** | **Overall (N = 452)** | **p-value[3]** |
| Diverticulitis, n (%)[1] | 452 | 98 (44%) | 30 (55%) | 76 (43%) | 204 (45%) | 0.318 | 582 | 91 (40%) | 38 (44%) | 112 (42%) | 241 (41%) | 0.778 |
| Haemorrhoids, n (%)[1] | 452 | 48 (22%) | 8 (15%) | 30 (17%) | 86 (19%) | 0.332 | 582 | 45 (20%) | 10 (11%) | 29 (11%) | 84 (14%) | **0.015** |
| Analfissure, n (%)[1] | 452 | 1 (0.5%) | 0 (0%) | 0 (0%) | 1 (0.2%) | >0.999 | 582 | 1 (0.4%) | 0 (0%) | 1 (0.4%) | 2 (0.3%) | >0.999 |
| Angiodysplasia, n (%)[1] | 452 | 6 (2.7%) | 1 (1.8%) | 2 (1.1%) | 9 (2.0%) | 0.453 | 582 | 9 (3.9%) | 3 (3.4%) | 6 (2.3%) | 18 (3.1%) | 0.522 |
| Rectum prolapse, n (%)[1] | 452 | 0 (0%) | 0 (0%) | 0 (0%) | 0 (0%) | >0.999 | 582 | 1 (0.4%) | 0 (0%) | 0 (0%) | 1 (0.2%) | 0.543 |
| IBD, n (%)[1] | 452 | 9 (4.1%) | 0 (0%) | 1 (0.6%) | 10 (2.2%) | **0.045** | 582 | 12 (5.2%) | 0 (0%) | 3 (1.1%) | 15 (2.6%) | **0.005** |
| Other GI-related symptoms, n (%)[1] | 452 | 4 (1.8%) | 2 (3.6%) | 1 (0.6%) | 7 (1.5%) | 0.161 | 582 | 9 (3.9%) | 0 (0%) | 6 (2.3%) | 15 (2.6%) | 0.13 |
| Antibiotics, n (%)[2] | 452 | 89 (40%) | 20 (36%) | 60 (34%) | 169 (37%) | 0.443 | 582 | 56 (24%) | 22 (25%) | 78 (29%) | 156 (27%) | 0.448 |
| Drugs indicating COPD, n (%)[2] | 452 | 42 (19%) | 14 (25%) | 33 (19%) | 89 (20%) | 0.517 | 582 | 26 (11%) | 14 (16%) | 46 (17%) | 86 (15%) | 0.166 |
| Drugs indicating CVD, n (%)[2] | 452 | 135 (61%) | 37 (67%) | 100 (57%) | 272 (60%) | 0.357 | 582 | 156 (68%) | 70 (80%) | 170 (64%) | 396 (68%) | **0.016** |
| Drugs indicating T2DM, n (%)[2] | 452 | 17 (7.7%) | 3 (5.5%) | 4 (2.3%) | 24 (5.3%) | **0.044** | 582 | 28 (12%) | 7 (8.0%) | 33 (12%) | 68 (12%) | 0.518 |

[1]Recorded by endoscopist during colonoscopy examination.
[2]Drug use was assessed using data from the Norwegian Prescription Registry, identifying individuals who had been prescribed medications indicative of common comorbidities.
[3]Statistical tests across colonoscopy groups, stratified on sex, were performed with Pearson's Chi-squared test; Fisher's exact test.

**Supplementary Table 4.** Odds ratios (ORs) and 95% confidence intervals (CIs) for presence of non-advanced neoplastic lesions[1] and advanced lesions[2] relative to controls by sex (n=1,034)[3].

| | **No neoplasia (n=450)** | **Non-advanced neoplastic lesions (n=142)** | | | **Advanced lesions (n=442)** | | |
|---|---|---|---|---|---|---|---|
| | n | n | OR (95% CI) | p-value | n | OR (95% CI) | p-value |
| **Female** | 221 | 55 | Ref. | | 176 | Ref. | |
| **Male** | | | | | | | |
| *Model 1* | 229 | 87 | 1.55 (1.05, 2.29) | **0.028** | 266 | 1.47 (1.12, 1.92) | **0.005** |
| *Model 2* | 229 | 87 | 1.50 (1.00, 2.26) | 0.052 | 266 | 1.46 (1.10, 1.93) | **0.009** |

[1]Includes any adenoma (adenomatous polyp) not fulfilling the criteria of being advanced.

[2]Includes advanced adenoma, defined as any adenoma with either villous histology (≥25% villous components), high-grade dysplasia or polyp size greater than or equal to 10 mm; advanced serrated lesions, defined as any serrated lesions with size ≥ 10 mm or dysplasia; and colorectal cancer, defined as presence of adenocarcinoma arising from the colon or rectum.

[3]Odds ratios (ORs) and 95% confidence intervals (CIs) are obtained from multinomial logistic regression analyses adjusting for the following covariates: Model 1: age (continuous) and screening centre (Moss, Bærum); model 2: model 1 covariates, employment status (employed, retired, other), marital status (not married/non-cohabiting, married/cohabiting), smoking status (never smoked, ever smoked) and the WCRF/AICR score (continuous).

**Supplementary Table 5**: Prevalence of previously associated CRC bacteria.

| | **Women** | | | | | **Men** | | | | |
|---|---|---|---|---|---|---|---|---|---|---|
| **Characteristic, n (%)** | **N** | **No adenoma (N = 221)** | **CRC (N = 31)** | **Overall (N = 252)** | **p-value[1]** | **N** | **No adenoma (N = 221)** | **CRC (N = 31)** | **Overall (N = 252)** | **p-value[1]** |
| *GGB9342_SGB14306* | 252 | 90 (41%) | 18 (58%) | 108 (43%) | 0.068 | 264 | 87 (38%) | 21 (60%) | 108 (41%) | **0.014** |
| *Alistipes ihumii* | 252 | 104 (47%) | 19 (61%) | 123 (49%) | 0.138 | 264 | 106 (46%) | 27 (77%) | 133 (50%) | **<0.001** |
| *Fusobacterium nucleatum* | 252 | 1 (0.5%) | 0 (0%) | 1 (0.4%) | >0.999 | 264 | 3 (1.3%) | 2 (5.7%) | 5 (1.9%) | 0.132 |
| *Porphyromonas somerae* | 252 | 3 (1.4%) | 0 (0%) | 3 (1.2%) | >0.999 | 264 | 3 (1.3%) | 4 (11%) | 7 (2.7%) | **0.007** |
| *Porphyromonas uenonis* | 252 | 4 (1.8%) | 0 (0%) | 4 (1.6%) | >0.999 | 264 | 1 (0.4%) | 2 (5.7%) | 3 (1.1%) | **0.047** |
| *Dialister pneumosintes* | 252 | 2 (0.9%) | 2 (6.5%) | 4 (1.6%) | 0.075 | 264 | 9 (3.9%) | 5 (14%) | 14 (5.3%) | 0.025 |
| *Gemella morbillorum* | 252 | 5 (2.3%) | 4 (13%) | 9 (3.6%) | **0.015** | 264 | 9 (3.9%) | 3 (8.6%) | 12 (4.5%) | 0.203 |
| *Fusobacterium nucleatum* | 252 | 2 (0.9%) | 2 (6.5%) | 4 (1.6%) | 0.075 | 264 | 3 (1.3%) | 1 (2.9%) | 4 (1.5%) | 0.436 |
| *Peptostreptococcus stomatis* | 252 | 4 (1.8%) | 4 (13%) | 8 (3.2%) | **0.009** | 264 | 9 (3.9%) | 4 (11%) | 13 (4.9%) | 0.077 |

[1]Fisher's exact test; Pearson's Chi-squared test

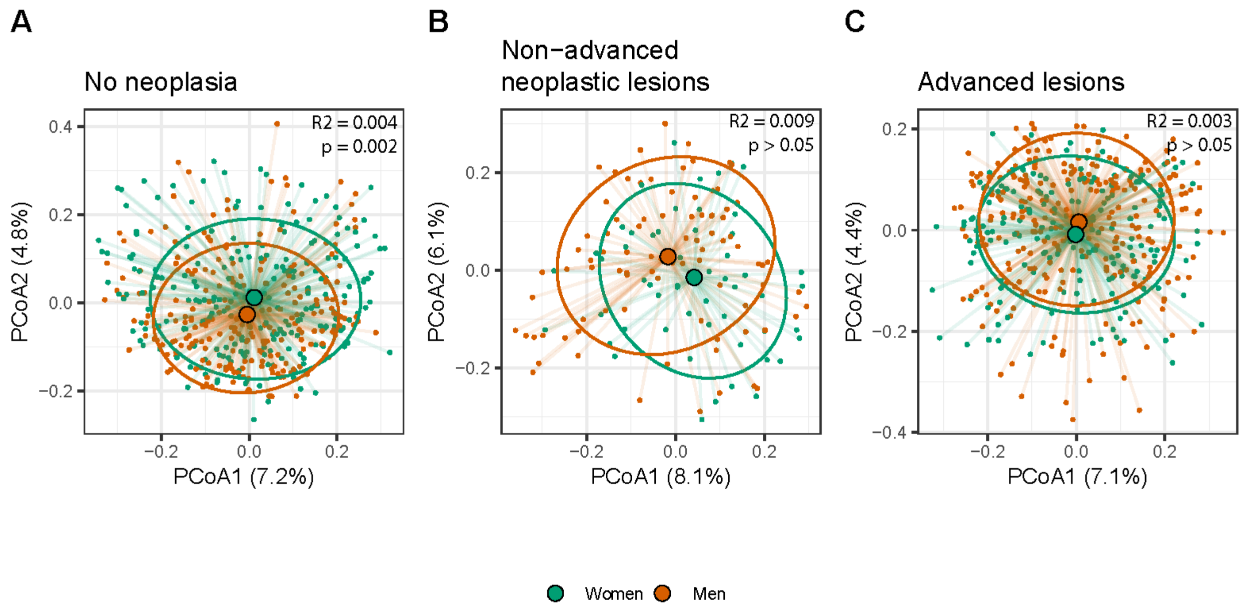


**Supplementary Figure 1.** Differences in β-diversity between women and men across screening findings A) no neoplasia, B) non-advanced neoplastic lesions, and C) advanced lesions. Analyses were adjusted for age (continuous), screening centre (Moss, Bærum), employment status (employed, retired, other), marital status (not married/non-cohabiting, married/cohabiting), smoking status (never smoked, ever smoked), and the WCRF/AICR score (continuous).

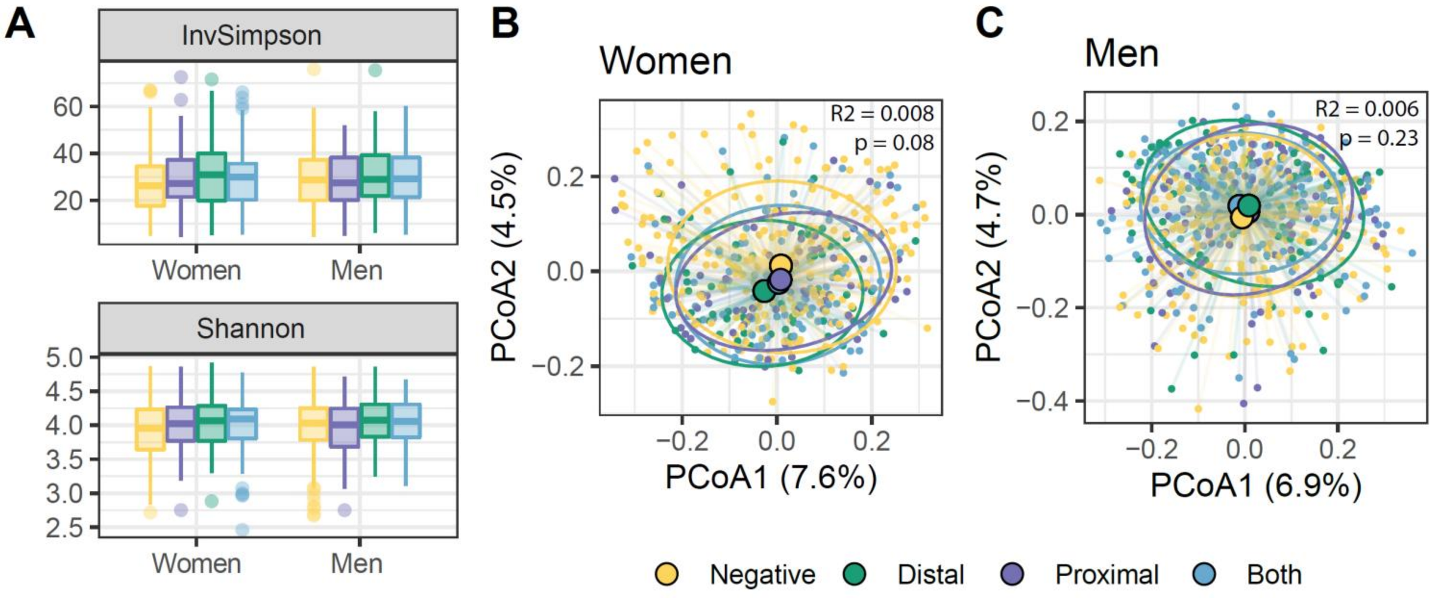


**Supplementary Figure 2.** Differences in β-diversity between women and men across screening findings A) no neoplasia, B) non-advanced neoplastic lesions, and C) advanced lesions. Analyses were adjusted for age (continuous), screening centre (Moss, Bærum), employment status (employed, retired, other), marital status (not married/non-cohabiting, married/cohabiting), smoking status (never smoked, ever smoked), and the WCRF/AICR score (continuous).
